# Compositional coding of individual finger movements in human posterior parietal cortex and motor cortex enables ten-finger decoding

**DOI:** 10.1101/2022.12.07.22283227

**Authors:** Charles Guan, Tyson Aflalo, Kelly Kadlec, Jorge Gámez de Leon, Emily R. Rosario, Ausaf Bari, Nader Pouratian, Richard A. Andersen

## Abstract

**Objective:** Enable neural control of individual prosthetic fingers for participants with upper-limb paralysis.

**Approach:** Two tetraplegic participants were each implanted with a 96-channel array in the left posterior parietal cortex (PPC). One of the participants was additionally implanted with a 96-channel array near the hand knob of the left motor cortex (MC). Across tens of sessions, we recorded neural activity while the participants attempted to move individual fingers of the right hand. Offline, we classified finger movements from neural firing rates using linear discriminant analysis (LDA) with cross-validation. The participants then used the neural classifier online to control individual fingers of a brain-machine interface (BMI). Finally, we characterized the neural representational geometry during individual finger movements of both hands.

**Main Results:** The two participants achieved 86% and 92% online accuracy during BMI control of the contralateral fingers (chance = 17%). Offline, a linear decoder achieved ten-finger decoding accuracies of 70% and 66% using respective PPC recordings and 75% using MC recordings (chance = 10%). A compositional code linked corresponding finger movements of the contralateral and ipsilateral hands.

**Significance:** This is the first study to decode both contralateral and ipsilateral finger movements from PPC. Online BMI control of contralateral fingers exceeded that of previous finger BMIs. PPC and MC signals can be used to control individual prosthetic fingers, which may contribute to a hand restoration strategy for people with tetraplegia.

## Introduction

Tetraplegic individuals identify hand function as a high-impact priority for improving their quality of life [1–3]. Neuroprosthetics research has enabled control of basic grasp shapes [4,5], an important step towards empowering paralyzed individuals to perform daily activities. However, these basic grasp templates constrain the range of motion and thus limit the usefulness of existing neural prosthetics.

The complexity of human motor behavior is largely enabled by our versatile, dexterous hands [6]. The human hand can weave intricate crafts, sign expressive languages, and fingerpick guitar solos. Even everyday manual behaviors, like turning a door handle, require volitional control over many degrees of freedom [7]. Indeed, humans can move individual fingers much more independently than other animals, including monkeys [8,9]. To better restore autonomy to people with tetraplegia, neural prosthetics would benefit from enabling dexterous finger control.

Cortical brain-machine interface (BMI) research has focused on control of computer cursors and robotic arms, rather than dexterous hand control. Foundational studies implemented continuous decoders for cursor control [10–13]. Leveraging this cursor control, [14,15] subsequently developed on-screen keyboard typing interfaces. [5,16–18] applied continuous decoding to arm control, with [16] controlling the user’s own muscles. Recent work has also decoded speech from sensorimotor cortex [19–22]. However, relatively few BMI studies have focused on hand control [23–28], and previous studies frequently combine the ring and little fingers or leave them out altogether. Individuated finger control would be useful for applications like keyboard typing or object manipulation.

Most motor BMIs record neural activity from the motor cortex (MC), although areas of the posterior parietal cortex (PPC) have also been used successfully for BMI control (for review, see [29]) of reaching [10,12,30] and grasping [4,22]. The PPC plays a central role in sensorimotor integration, with regions of PPC representing visual stimulus locations [31], eye movements [32], task context [33], planned reaches [34], and object grasping [35,36]. PPC uses partially mixed selectivity to simultaneously encode many motor variables [37], which can be useful for versatile neural decoding.

Despite PPC’s clearly demonstrated role in dexterous grasping [6,36,38], less is known about PPC responses during individual finger movements. With fMRI, lesion, and anatomical evidence situating primary motor cortex as core to fine finger movements (for review, see [6]), most electrophysiological studies of finger movements have focused on the primary motor (M1) and primary somatosensory cortex (S1) [24,25,28,39–42]. Nevertheless, non-human primate mapping studies [43] and stimulation studies [44,45] have identified PPC sub-regions that are likely involved in fine finger movements. These results imply that fine finger movements are supported by a broad neuronal network, which should be investigated to improve dexterous BMI control.

Here, we recorded intracortical activity from the PPC of two tetraplegic participants while they attempted to press individual fingers. Across task contexts, we could classify individual finger movements during planning and execution periods. We connected this neural decoder to drive a neural prosthetic hand, with accuracies exceeding recent intracortical BMI studies [27,46]. Furthermore, we characterize both the neural tuning and representational geometry [47] during finger movements of both hands. The neural code was composed of separable laterality and finger components, leading to finger representations that were simultaneously discriminable and similar across contralateral/ipsilateral pairs of fingers. These findings contribute to the understanding of human hand movements and advance the development of hand neuroprosthetics for people with paralysis.

## Methods

### Study participants

Experiments were conducted with volunteer participants enrolled in a brain-machine interface (BMI) clinical study (ClinicalTrials.gov Identifier: NCT01958086). All procedures were approved by the respective Institutional Review Boards of California Institute of Technology, Casa Colina Hospital and Centers for Healthcare, and University of California, Los Angeles.

Participant N is a right-handed, tetraplegic woman. Approximately 10 years before this study, she sustained an AIS-A spinal cord injury at cervical level C3-C4. N can move her deltoids and above, but she cannot move or feel her hands.

Participant J is a right-handed, tetraplegic man. Approximately 3 years before this study, he sustained a spinal cord injury at cervical level C4-C5. He has residual movement in his upper arms, but he cannot move or feel his hands.

Each participant consented to this study after understanding the nature, objectives, and potential risks.

### Tasks

#### Alternating-cues finger press task with delay

Each participant performed an instructed-delay finger movement task (Figure 1). They were seated in front of a computer monitor display, with their hands prone on a flat surface. Each trial began with a cue specifying a finger of the right hand. The finger cue then disappeared during a delay period. A condition-invariant go-icon appeared, instructing the participant to attempt to press the cued finger as though pressing a key on a keyboard. This instructed-delay task format temporally separates the visual stimulus from the planning and execution epochs.

**Figure 1.**
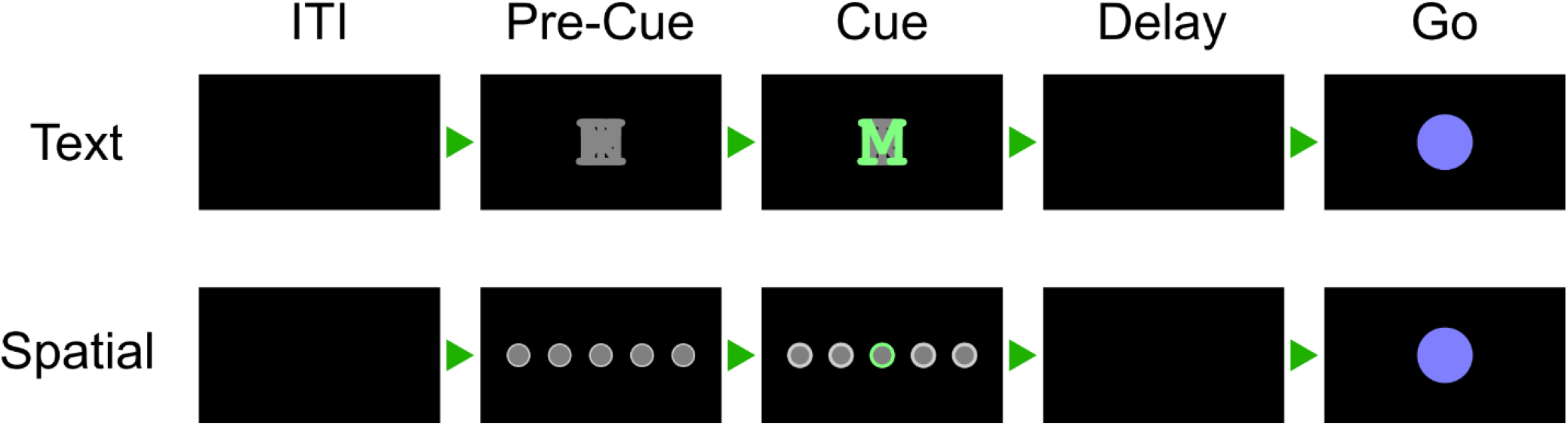
Alternating-cues, instructed-delay finger press task. Trial structure. Each rectangle represents the computer monitor display at each phase. Two cue variants, text and spatial, were trial-interleaved. In the spatial variant, the location of the highlighted circle corresponded to the cued finger. Trials without a highlighted circle indicated a No-Go cue. In the text variant, a highlighted letter (for example, “M” for the middle finger) cued each finger. In both variants, the finger cue disappeared before the movement phase (Go) to separate planning and execution periods. Phase durations are listed in Supplementary Table 1.

Supplementary Table 1 documents the phase durations for each task, and Supplementary Table 2 lists the date ranges for each task.

Some regions of the posterior parietal cortex (PPC) are modulated by non-motor variables like visual stimulus location [31] and task context [33]. To ensure that the recorded neural signals reflected movement type (rather than, e.g., visual memory), we varied the cueing method between runs (Figure 1). In the Spatial-Cue variant, five circles corresponded to the five fingers. In the Text-Cue variant, the finger cue was a letter abbreviation. A brief Pre-Cue phase in each trial indicated what cue variant the trial would be.

#### Finger press task with randomized cue location (reaction-time)

Letters, corresponding to each movement type, were arranged in a 3 × 4 grid across the screen. Each grid consisted of two repetitions each of T (thumb), I (index), M (middle), R (ring), P (pinky), and X (No-Go). Letters were arranged in a random order to dissociate eye gaze signals from movement representations. On each trial, a single letter cue was indicated with a crosshairs symbol, which was jittered to minimize systematic effects of letter occlusion. Each cue was selected once (for a total of 12 trials) before the screen was updated to a new arrangement. Each run-block consisted of 4 screens for a total of 48 trials.

On each trial, the participant was instructed to immediately saccade to the cued target and fixate, then attempt to press the corresponding finger of the right hand. A trained classifier (<u>Brain-machine interface (BMI) calibration</u>) decoded the finger movement from neural signals (<u>Online BMI discrete control</u>) and displayed the classified finger movement 1.5 seconds after the start of the trial. The participant pressed the instructed finger and fixated on the cue until the visual classification feedback was shown.

Data from participant N performing this task was previously analyzed in [46]. Data from participant J have not been reported previously. During 3 sessions, participant J also performed this task using his left hand.

#### Ten-finger press task

Each participant also performed an instructed-delay finger press task with fingers from both hands. The task was like the Text-Cue variant of the Alternating-cues finger press task with delay, except without a Pre-Cue phase. All ten fingers were interleaved in trials within the same run-block (Figure 3). Phase durations are documented in Supplementary Table 1.

**Figure 2.**
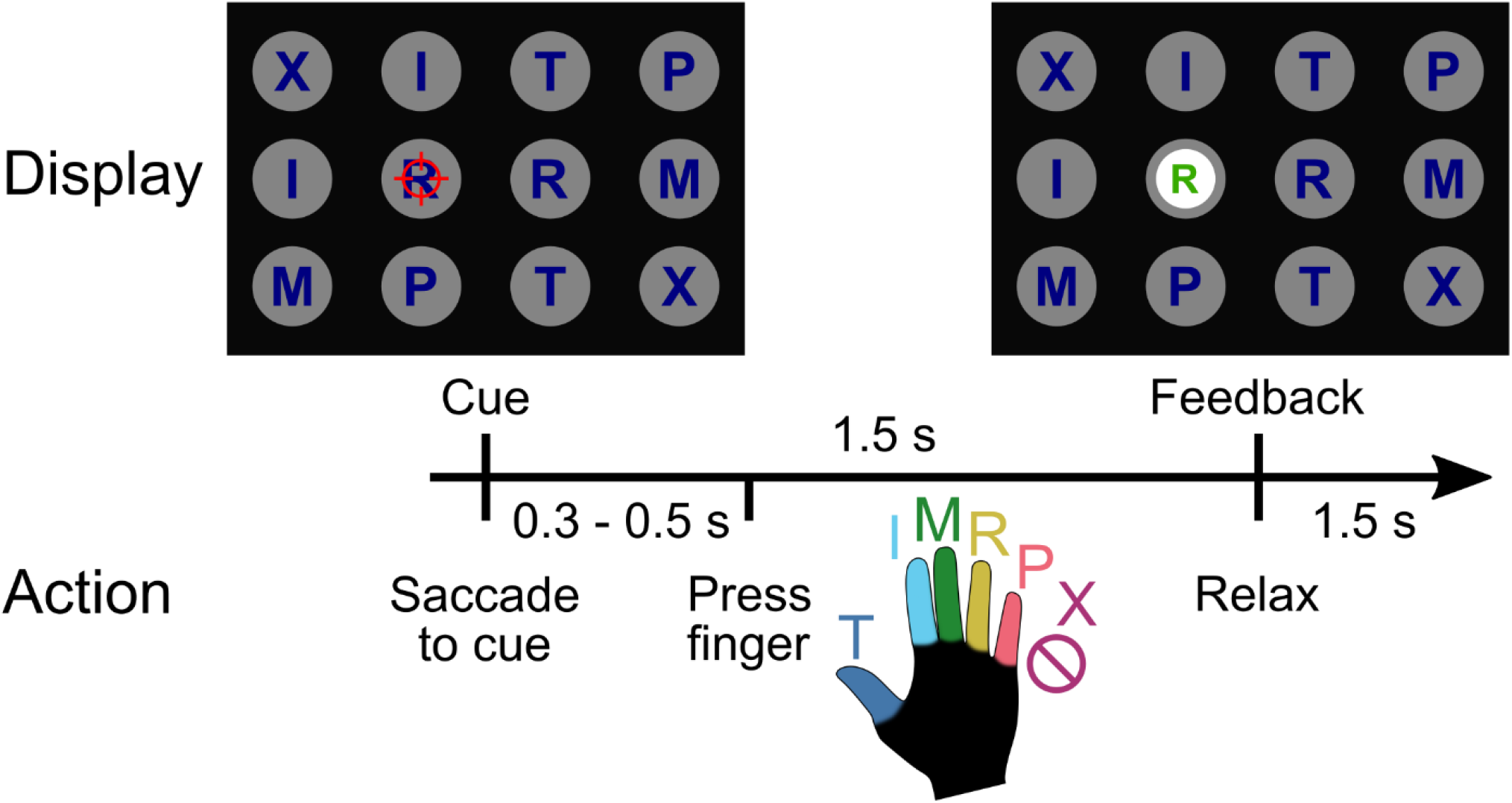
Reaction-time finger-press task with randomized cue location. Figure adapted from [46] (CC BY-NC 4.0). Main finger press task. When a letter was cued by the red crosshair, the participant looked at the cue and immediately attempted to flex the corresponding digit of the right (contralateral) hand. We included a null condition “X,” during which the participant looked at the target but did not move their fingers. Visual feedback indicated the decoded finger 1.5 seconds after cue presentation. To randomize the saccade location, cues were located on a grid (3 rows, 4 columns) in a pseudorandom order. The red crosshair was jittered to minimize visual occlusion.

**Figure 3.**
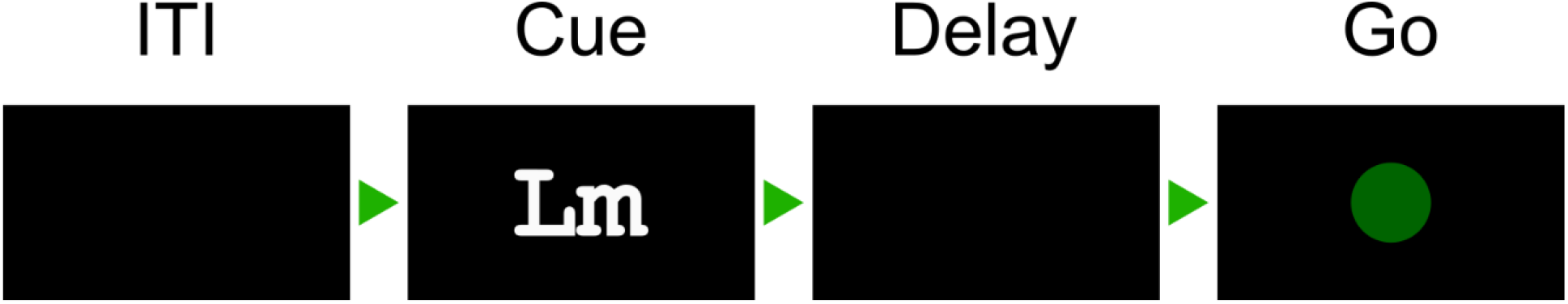
Text-cued finger movement task with instructed-delay. Trial structure. Text cues indicate the hand (“R” or “L”) and the finger (e.g., “m” for middle finger). A delay period separates the Cue phase from a condition-independent Go display.

### Implant location

Participant N was implanted with two 96-channel NeuroPort Utah electrode arrays 6 years after injury (about 4 years before this study). The implant locations were determined using anatomical priors and preoperative functional magnetic resonance imaging (fMRI) [46]. One array (denoted N-PPC) was implanted over the hand/limb region of PPC at the junction of the intraparietal sulcus (IPS) with the postcentral sulcus (PCS). This region is thought to be involved in the planning of grasp movements [4,36,48]. In this report, we refer to this brain area as PC-IP (postcentral-intraparietal), although it is sometimes also referred to as the anterior intraparietal sulcus (aIPS) region [49]. A second array was in Brodmann’s area (BA) 5d. In the weeks following implantation, it was found that the BA 5d array did not function, so only the PC-IP array was used in this study.

Participant J was implanted with two 96-channel NeuroPort Utah electrode arrays about 20 months after injury (about 35 months before this study). The first array (denoted J-PPC) was implanted in the superior parietal lobule (SPL) of the left PPC. The second array (denoted J-MC) was implanted near the hand knob of the left motor cortex (MC) (Supplementary Figure 1). PPC and MC activity were recorded simultaneously.

### Neural signal recording and preprocessing

Neural signals were acquired, amplified, bandpass-filtered (0.3 Hz - 7.5 kHz) and digitized (30 kHz, 16-bits/sample) from the electrodes using NeuroPort Neural Signal Processors (NSP) (Blackrock Microsystems Inc.).

Action potentials (spikes) were detected by high-pass filtering (250Hz cut-off) the full-bandwidth signal, then thresholding at −3.5 times the root-mean-square (RMS) voltage of the respective electrode. Although one or more source neurons may generate threshold crossings, we used raw threshold crossings for online control and only sorted spikes for offline analyses. Single neurons were identified using the k-medoids clustering method. We used the gap criteria [50] to determine the total number of waveform clusters. Clustering was performed on the first n ∈ {2, 3, 4} principal components, where n was selected to account for 95% of waveform variance.

### Feature Extraction

Except when otherwise specified, we used a 500-millisecond (ms) window of neural activity to calculate firing rates (counted spikes divided by the window duration). The firing rate was then used as the input features to each analysis or classification model. Neurons with an average firing rate less than 0.5 Hz were excluded from all analyses.

Behavioral epochs: the movement execution (“Go” or “move”) analysis window was defined as the 500-ms window starting 200 ms after the Go cue. For applicable tasks, the movement planning (“Delay” or “plan”) analysis window was defined as the 500-ms window starting 200 ms after the Delay screen. The Cue analysis window was defined as the 500-ms window starting 200 ms after the Cue screen. The intertrial interval (ITI) analysis window was defined as the last 500 ms of the ITI phase.

### Single-neuron selectivity for finger movements

In the section “Single-neuron modulation to individual finger presses,” we used a one-way ANOVA to determine whether neurons distinguished firing rates between conditions. A neuron was considered discriminative if p < 0.05 after false discovery rate (FDR) correction for multiple comparisons using the Benjamini–Hochberg procedure; we also denoted this FDR-adjusted p-value as *q*. We corrected for *m*=*N* comparisons, where N is the number of neurons for each participant. Following Cohen’s rules of thumb [51], we denoted the ANOVA effect size as “large” if η^2^ > 0.14. As the ANOVA post hoc test, we used Dunnett’s multiple comparison test [52] to determine which fingers had significantly different firing rates than the No-Go baseline.

To quantify the effect size of firing-rate changes against the No-Go baseline (Figure 4a), we used Hedges’ g, which is similar to Cohen’s d but bias-corrected for small sample sizes. We calculated and visualized Hedges’ g values using the Data Analysis using Bootstrap-Coupled Estimation Python library [53].

**Figure 4.**
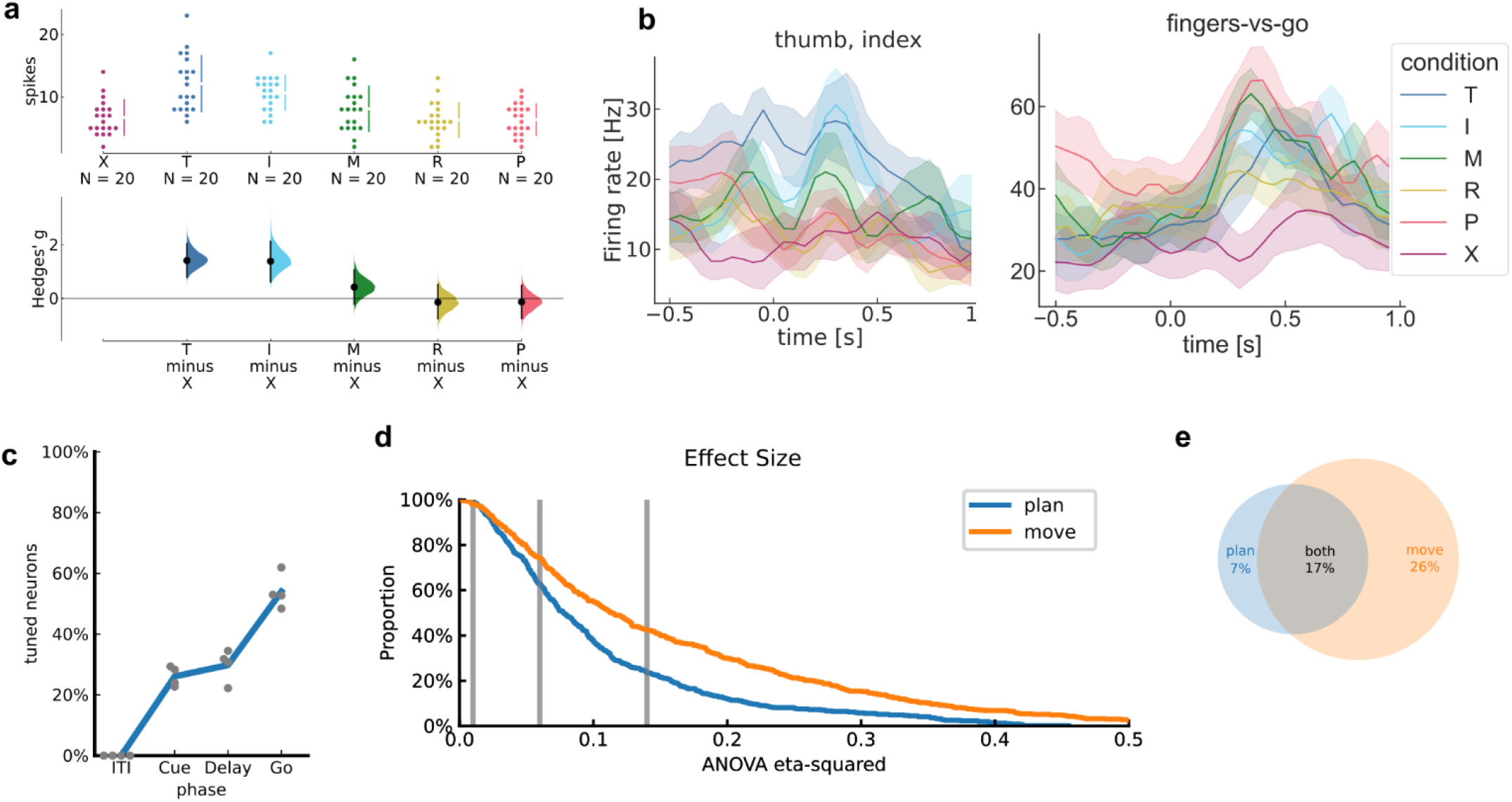
PPC single neurons discriminate between attempted finger movements. a) Single-trial firing rates for an example N-PPC neuron during movements of different fingers. (top) Markers correspond to the firing rate during each trial (N=120 across 6 conditions). Gapped vertical lines to the right of markers indicate +/- S.D., and each gap indicates the condition mean. (bottom) Firing rates during thumb (T) and index (I) presses were higher than the No-go (X) baseline. Vertical bars indicate bootstrap 95% confidence intervals (CI) of the effect size versus No-go baseline. Half-violin plots indicate bootstrap distributions. b) Mean smoothed firing rates for each finger movement for two example N-PPC neurons. Shaded areas indicate 95% CI. c) Percentage of N-PPC neurons that discriminated between finger movements in each analysis window (q < 0.05, FDR-corrected for 466 neurons). Line (blue) indicates mean across sessions. Markers (gray) indicate individual sessions. d) Complementary empirical cumulative distribution function (cECDF) visualizing the proportion of N-PPC neurons with ANOVA effect sizes (η^2^) above the corresponding x-axis value. Vertical lines (gray) indicate Cohen’s thresholds [51] for small (η^2^=0.01), medium (η^2^=0.06), and large (η^2^=0.14) effect sizes. e) Overlap of N-PPC neurons that modulated significantly (q < 0.05) with large effect sizes (η^2^ > 0.14) during movement preparation and movement execution.

For visual simplicity, we pooled neurons across sessions when calculating and visualizing single-neuron metrics (percentage selective, number of fingers discriminable from No-Go, empirical cumulative distribution functions).

To visualize firing rates, spike rasters were smoothed with a Gaussian kernel (50-ms standard-deviation [S.D.]), then averaged across trials to create a peristimulus time histogram (PSTH).

### Offline classification with cross-validation

We trained a separate linear classifier for each session to predict finger movements from the neural features. We used diagonal-covariance linear discriminant analysis (diagonal LDA) [54]; Diagonal LDA is equivalent to Gaussian Naive Bayes (GNB) when GNB shares a single covariance matrix across classes.

For offline classification and parameter sweeps, we used stratified K-Folds cross-validation (with K = 8) to estimate the generalization error. Reported classification metrics correspond to cross-validated performance on the evaluation sets. Results were aggregated over sessions by pooling trials. Across-session standard deviations of classification accuracy are weighted by the number of trials in each session.

Learning curves (Figure 5b) were generated by using subsets of the training set during each Stratified K-Fold split. Neuron-dropping curves (Figure 5c) were generated by stacking neurons across sessions, then evaluating performance with random subsets of neurons. Window duration sweeps (Figure 5d) varied the size of the firing-rate estimation window while fixing the start time at 200ms after the Go cue. Neural decode time-courses (Figure 5e) used 500ms bins centered at different times of the trial.

**Figure 5.**
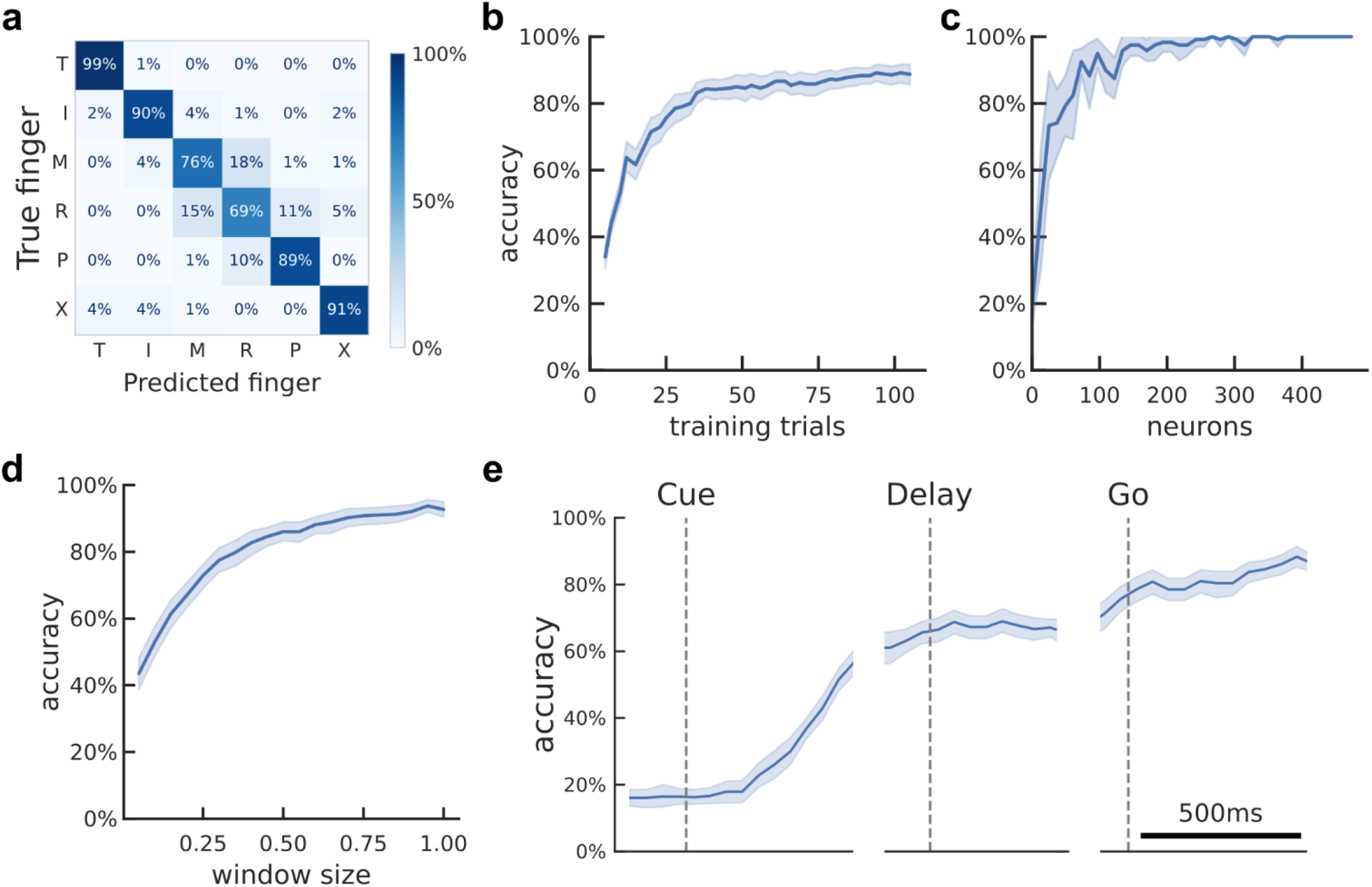
Offline classification of finger movement execution from population activity. a) Cross-validated confusion matrix for classifying finger movement from N-PPC neural activity. 86% accuracy, 480 trials over 4 sessions. b) Learning curve showing cross-validated accuracy as a function of the training dataset size. About 40 trials (less than 7 trials per finger) are needed to achieve 80% accuracy. Shaded area indicates 95% CI over folds/sessions. c) Neuron-dropping curve (NDC) showing cross-validated accuracy as a function of recorded neurons. Neurons were aggregated across sessions. About 70 neurons are needed to achieve 80% accuracy. d) Hyperparameter sweep showing cross-validated classification accuracy as a function of decode window size. Input features were the average firing rates in the window [200ms, 200ms + *window size*] after Go-cue. Window durations of about 350ms are necessary to achieve 80% accuracy. e) Cross-validated classification accuracy across the trial duration (500-ms sliding window).

### Online brain-machine interface (BMI) discrete control

Each BMI control session started with a run of the open-loop calibration task. For participant N, this was the Alternating-cues finger press task, modified to not have a delay. For participant J, this was the finger press task with randomized cue location, without classifier output.

The neural activity and finger movements from the calibration task served as training data for the online BMI classification model. Neural features were composed of the threshold crossing rates of each electrode during a 1-second window. Electrodes with mean firing rates less than 1 Hz were excluded to minimize sensitivity to discretization. The window start-time was chosen to maximize the cross-validated classification accuracy on the calibration task. The classifier was then re-trained using all data from the calibration task (without cross-validation), using threshold crossing rates from the selected window. This classifier was then used to decode attempted movements in the BMI task.

During online control of the finger grid task, the classifier predicted a single finger movement for each trial. Input neural features consisted of the threshold crossing rates from each electrode in the time window [0.5, 1.5] seconds after cue presentation.

As a proof-of-concept, we also connected the classifier output to the fingers of a robot hand (not shown in preprint). On each trial, a screen cue instructed the participant which finger to press. The BMI classifier predicted each finger movement from the neural features and then moved the corresponding finger on the robotic hand.

### Neural distance between fingers

We quantified the neural activity differences between finger movements using the cross-validated (squared) Mahalanobis distance [55]. The Mahalanobis distance is a continuous, non-saturating analogue of LDA classification accuracy [56]. Cross-validation removes the positive bias of standard distance metrics, such that 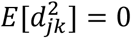 when two activity patterns are statistically identical.

To calculate population distances, we used the representational similarity analysis Python toolbox [57]. The toolbox slightly modifies the cross-validated Mahalanobis equation, incorporating the noise covariances of both folds to improve robustness:

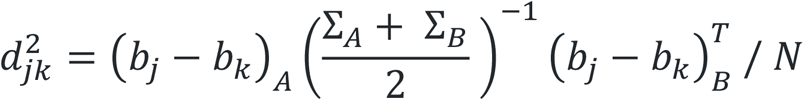

where *A* and *B* indicate independent partitions of the trials, Σ is the noise covariance matrix, (*b*_*j*_, *b*_*k*_) are the firing rate vectors for conditions (*j, k*) stacked across trials, and *N* normalizes for the number of neurons. The units of 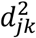 are *unitless*^2^/*neuron*.

### Shared representations across hands

To quantify whether finger representations were similar across hands, we compared the pairwise distances between matching finger pairs and the pairwise distances between non-matching finger pairs (Figure 8b). We denoted a finger pair as matching if the hands differed and the finger-types were the same ([Lt, Rt], [Li, Ri], [Lm, Rm], [Lr, Rr], [Lp, Rp]). We denoted a finger pair as non-matching if the hands differed and the finger-types also differed ([Lt, Ri], [Lt, Rm], [Lt, Rr], [Lt, Rp], [Li, Rt], [Li, Rm], etc.). We described a neural population as sharing representations across hands if the average distance between matching finger pairs was smaller than the average distance between non-matching finger pairs.

### Compositionality of finger representations

Compositional coding refers to representations that can be constructed by combining and recombining basic components [58,59]. We assessed whether finger representations could be linearly decomposed into the sum of finger-type and laterality components.

We first visualized the representational geometry in Figure 8d using 2-D multidimensional scaling (MDS). MDS projects the movement conditions into a low-dimensional space that preserves pairwise distances (Figure 8a) as well as possible. We performed MDS on data from individual sessions and then used Generalized Procrustes Analysis (GPA) with scaling to normalize and align MDS projections across sessions. In the N-PPC MDS plot, ellipses show standard error (S.E.) across sessions. The J-PPC and J-MC MDS plots show the condition means without any S.E. ellipses, because the 2 sessions with participant J are not sufficient to estimate the S.E.

To assess compositionality, we used leave-one-group-out cross-validation to determine whether hand- and finger-dimensions generalize to left-out movements (Supplementary Figure 8). If finger representations are compositional, then hand classifiers (left vs. right) should generalize when trained on a subset of finger types and evaluated on left-out fingers. Additionally, finger-type classifiers should generalize when trained on one hand and tested on the other hand (Figure 8e). This metric is sometimes called cross-condition generalization performance (CCGP) [60,61]. We pooled neurons across sessions (N: 10 sessions; J: 2) into a pseudo-population. We used a permutation test to assess whether CCGP was significantly above chance, shuffling the labels repeatedly (N=1001) to generate a null distribution. Standard, within-condition decoding accuracy provided an upper bound on CCGP. Reaching this upper bound implies perfect compositionality.

## Results

### Single-neuron modulation to individual finger presses

We first sought to determine whether PPC single neurons discriminate between individual finger movements. We quantified single-neuron modulation to attempted finger presses of the right (contralateral to the implant) hand while the participant performed the Alternating-cues finger press task with delay (participant N: 120 trials per session for 4 sessions; participant J: 112 trials per session [min: 96; max: 120] for 3 sessions). We recorded 118 neurons per session (min: 111; max: 128) over 4 sessions from N-PPC, 103 neurons per session (min: 92; max: 116) over 3 sessions from J-PPC, and 93 neurons per session (min: 90; max: 95) from J-MC. For each neuron, we calculated firing rates during the attempted movement period and compared firing rates across conditions (Figure 4a, Supplementary Figure 2, Supplementary Figure 3).

Similar to results from finger studies of the motor cortex hand area [42,62], PPC neurons were not anatomically segregated by finger selectivity. A large portion of neurons (N-PPC: 54%; J-PPC: 30%; J-MC: 78%; Figure 4c) discriminated firing rates across conditions (q < 0.05), and selective neurons were often selective for multiple finger movements (mean number of significant fingers, N-PPC: 2.1; J-PPC: 1.9; J-MC: 2.7). Moreover, many neurons discriminated between finger conditions with large effect sizes (percentage of neurons with η^2^ > 0.14, N-PPC: 40%; J-PPC: 25%; J-MC: 64%; Figure 4d-e).

We also quantified single-neuron modulation during movement preparation. Preparatory activity discriminated between finger conditions with reasonable effect sizes (Figure 4d). Consistent with reaching studies of PPC [10], slightly fewer PPC neurons had strong tuning (q < 0.05 and η^2^ > 0.14) during movement preparation (N-PPC: 24%; J-PPC: 23%) than during movement execution (N-PPC: 43%; J-PPC: 24%) (Figure 4e).

### Classifying finger presses from neural activity

Since single neurons were tuned to finger movements, we evaluated whether attempted finger movements could be classified (offline) from the population neural activity. Using data from the same task, we trained linear classifiers and assessed finger classification accuracy on held-out trials using cross-validation (Methods). Classification accuracies substantially exceeded chance (N-PPC: 86%; J-PPC: 64%; J-MC: 84%; chance: 17%). The majority (N-PPC: 75%; J-PPC: 42%; J-MC: 67%) of errors misclassified an adjacent finger (Figure 5a, Supplementary Figure 4, Supplementary Figure 5).

Classification accuracy can depend on the neural signal quality and prediction window. To better understand how finger classification varies over dataset and classifier parameters, we quantified cross-validated accuracy across different training dataset sizes, neuron counts, and window durations (Figure 5b-d, Supplementary Figure 4, Supplementary Figure 5).

Cross-validated accuracy increased with more training data, reaching 80% accuracy when training on about 40 trials (2.7 minutes) for N-PPC. Higher neuron counts provide more finger information and thus improved classification accuracy, reaching 80% accuracy at about 70 neurons for N-PPC. These results indicate that a single electrode array in PPC provides sufficient information to control a discrete finger-press prosthetic.

Accuracy also increased when using longer window durations, reaching 80% at durations above 350ms. Longer window durations average out firing rates and thereby reduce the impact of measurement noise and behavioral variability on classification, but they directly mandate longer control delays. In some cases, it may be useful to minimize BMI control latency even at the expense of accuracy [63].

Finger movements could also be decoded from PPC during the planning period (Figure 5e)), although classification accuracy was lower (N-PPC: 66%; J-PPC: 61%; chance: 17%) than during movement execution.

### Brain-machine interface control of finger movements

We next mapped neural activity to finger movements to control an online finger BMI, where our participants would tap each finger and their attempted movement would be decoded. For this section, we replicated a usage scenario where a prosthetic user could decide to move a finger and immediately execute the movement, without needing a delay period.

We started each session with an open-loop calibration task where the participant attempted to press fingers according to visual cues (Methods). Using only a short calibration period (8 repetitions per condition, totaling about 2.5 minutes), each participant was able to use a classifier to accurately control individual fingers of the BMI.

The confusion matrix for participant N (Figure 6a) shows that she achieved high online control accuracies (86%; chance: 17%). These finger representations were robust across contexts and could be used in a range of environments. In one session, participant N used the BMI to control the fingers of a robotic hand (not shown in preprint).

**Figure 6.**
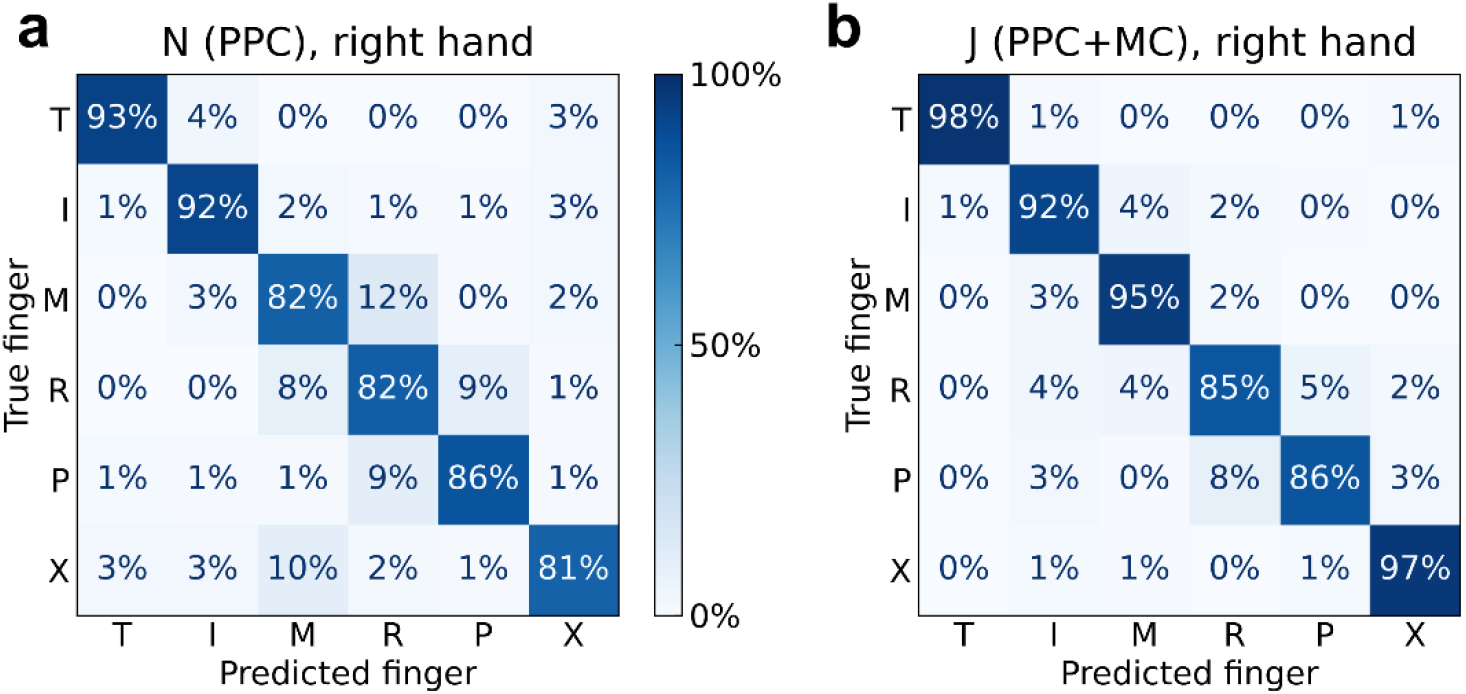
Online BMI classification of individual finger movements. a) Confusion matrix for participant N (PPC), right-hand finger presses. 86% accuracy, 4016 trials over 9 sessions. Reprinted from [46] (CC BY-NC 4.0). b) Confusion matrix for participant J (PPC+MC), right-hand finger presses. 92% accuracy, 1488 trials over 8 sessions.

Participant J achieved even higher accuracies during BMI control (92%; chance: 17%) (Figure 6b). However, we note that participant J’s BMI decoder used threshold crossings from both MC and PPC electrode arrays, thus doubling the number of electrodes compared to participant N. While we cannot retrospectively replicate the BMI experiment with an isolated array, we can approximate the results by training the same classification algorithm on early runs, using recordings only from a single array; we can then apply this classifier to the subsequent test trials (accuracy, J-PPC: 83%; J-MC: 87%; chance: 17%; Supplementary Figure 6).

On separate runs, participant J also performed this task with his left hand. Supplementary Figure 7 shows the corresponding confusion matrix.

### Decoding individual finger presses of either hand

We next investigated whether both contralateral and ipsilateral finger movements could be classified from a single array. Cerebral hemispheres primarily control movement on the opposite side of the body, and we have only implanted electrode arrays in each participant’s left hemisphere. However, the ability to classify movements of both sides would reduce the number of implants necessary for bilateral BMI applications.

We examined PPC neural activity patterns during interleaved, attempted finger presses of the contralateral (right) and ipsilateral (left) hands (Methods; participant N: 100 trials / session for 10 sessions; participant J: 100 trials / session for 2 sessions). We recorded 111 neurons per session (min: 102; max: 119) from N-PPC, 160 neurons per session (min: 159; max: 160) from J-PPC, and 130 neurons per session (min: 120; max: 130) from J-MC. Similar to the contralateral-only results, most neurons (N-PPC: 66%; J-PPC: 57%; J-MC: 78%) discriminated firing rates across fingers (q < 0.05).

We then evaluated whether these signals could be used for a neural prosthetic by classifying (offline) the attempted finger movement from the population neural activity. A linear classifier (Methods) was able to discriminate between all ten fingers (cross-validated classification accuracy, N-PPC: 70%; J-PPC: 66%; J-MC: 75%; chance: 10%). The majority (N-PPC: 76%; J-PPC: 66%; J-MC: 68%) of classification errors were adjacent-finger-confusion or matching-across-hand-confusion (Figure 7c-e).

**Figure 7.**
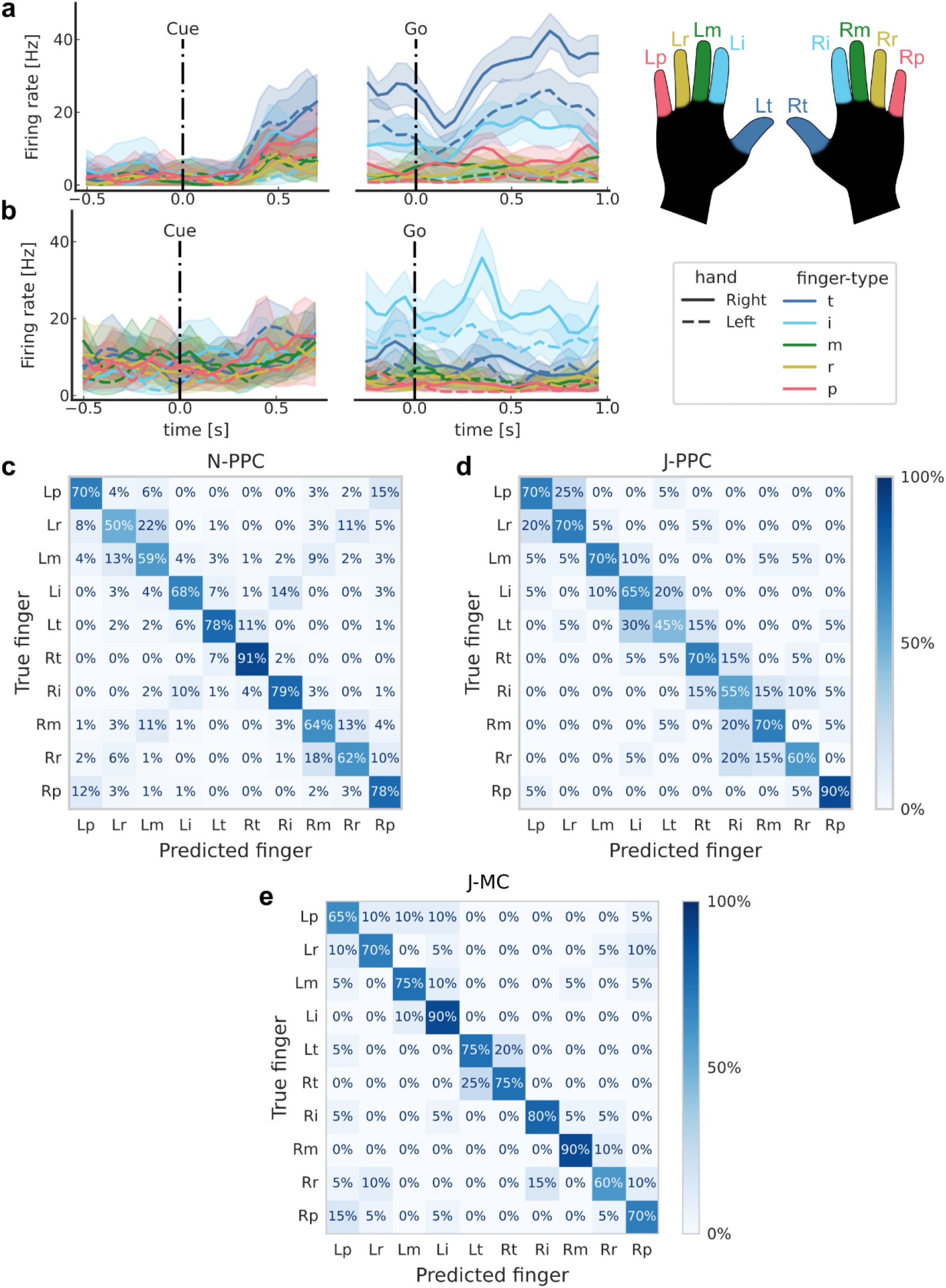
Classifying finger presses of both hands. a) Mean firing rates for each finger movement for an example N-PPC neuron, which increases its firing rate for thumb movements. Shaded areas indicate 95% confidence intervals (CI). b) Same as (a) for a second example N-PPC neuron, which increases it firing rate for index movements. c) Cross-validated confusion matrix for classifying right- and left-hand finger movements from N-PPC neural activity. 70% accuracy, 1000 trials over 10 sessions. d) Same as (c) using recordings from J-PPC. 66% accuracy, 200 trials over 2 sessions. e) Same as (c) using recordings from J-MC. 75% accuracy, 200 trials over 2 sessions.

### Compositional representation of finger and laterality

To characterize how PPC simultaneously represents contralateral and ipsilateral finger movements, we calculated the cross-validated neural distances between pairs of movement conditions. Figure 8a visualizes these distances in a representational dissimilarity matrix (RDM) [47] that is row- and column-indexed by the movement condition. Visual inspection shows that neural distances are small between right/left pairs of fingers (anti-diagonal of Figure 8a), suggesting that movement representations are partially shared across hands. On average, matching right/left finger pairs were 1.56 distance-units (95% CI: [1.33, 1.78], Figure 8b) closer to each other than non-matching fingers were. Matching fingers were also represented more similarly than non-matching fingers in J-MC (mean difference: 4.30, 95% CI: [2.74, 5.46]), but this result was not conclusive in J-PPC (mean difference: 0.27, 95% CI: [-0.17, 0.64]).

**Figure 8.**
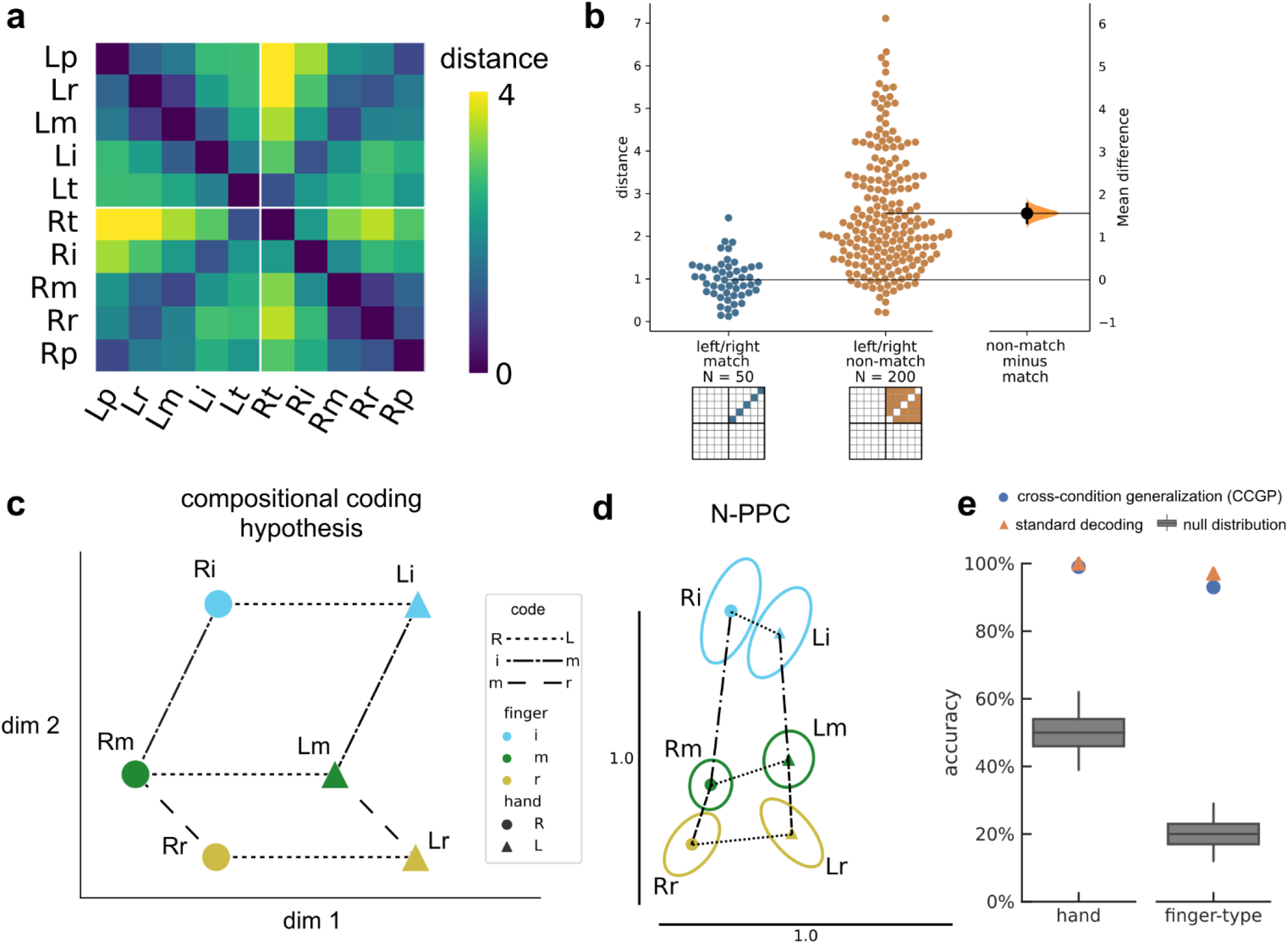
Representational geometry of contralateral and ipsilateral finger movements. a) Cross-validated squared Mahalanobis distances between N-PPC activity patterns during the contralateral/ipsilateral finger press task. Distances were averaged over the 10 sessions. b) Non-matching (different finger-type, different hand) finger pairs have larger distances than matching (same finger-type, different hand) finger pairs. Each circle is one element of the dissimilarity matrix of an individual session, aggregated across 10 sessions. c) Example schematic of perfect compositionality along hand and finger-type components. Line styles indicate groups of parallel, identical vectors. A compositional code can be represented as the linear summation of simple building blocks. For example, the Rm population activity can be constructed from the summation: Li + left->right + index->middle. For visual clarity, figure only shows three finger-types (index, middle, ring). d) Representational geometry of finger movements corresponding to N-PPC distances (a), visualized in 2-D using multidimensional scaling (MDS). We used Generalized Procrustes analysis (with scaling) to align across 10 sessions. Ellipses show standard error (S.E.) across sessions. Scale bars shown. Vectors with matching line-styles match each other, suggesting that the neural code is compositional. e) Linear decoders generalized (Supplementary Figure 8) across finger-type to classify hand (left) and across hand to classify finger-type (right) (p < 0.001, permutation test), indicating that movement representations were compositional across finger-type and hand dimensions.

What representational geometry allows downstream readout of all ten fingers (Figure 7) while sharing information across hands (Figure 8b)? Studies of human motor cortex [64–67] have also found correlated representations across sides, with [65] synthesizing the results under the framework of compositionality. Compositional codes refer to neural representations that can be constructed as the combination of simple building blocks [59]. [65] found that MC movement representations could be decomposed into simpler components: laterality, arm-versus-leg, and motion pattern.

Do laterality and finger-type also form a compositional code in PPC and MC? Perfect additive compositionality (Figure 8c) implies that the vector between neural representations is simply the summation of the vectors between their respective components. For example, the vector Lm->Ri can be decomposed into generic left->right and middle->index vectors. Geometrically, these generic vectors would form parallelograms between relevant groups of conditions (Figure 8c) [68]. Another interpretation is that compositional population activity would have a consistent hand subspace and a consistent finger-type subspace, although these subspaces could be non-orthogonal.

We used 2-D multidimensional scaling to visualize the geometric relationship between N-PPC finger representations (Figure 8d), limiting to the index, middle, and ring fingers for visual clarity. We found that inter-finger vectors were similar across hands, with the index finger relatively distinct from the middle and ring fingers, consistent with previous studies of contralateral finger movements [69,70]. Additionally, the left->right vector appeared identical across all matching left/right finger pairs.

Compositional coding allows generalization across the axes of the simpler building blocks. Since individual left->right vectors are nearly identical to each other, linear decoders trained to differentiate left-vs-right on a subset of finger types (Lt-vs-Rt; Li-vs-Ri, Lm-vs-Rm, Lr-vs-Rr) should generalize to held-out, hypothetically equivalent vectors (Lp-vs-Rp) (Supplementary Figure 8). Consistent with this hypothesis, cross-condition hand-decoding generalization performance (hand CCGP) was significantly above chance (p < 0.001, permutation test). Furthermore, hand CCGP was close to the standard decoding accuracy evaluated using cross-validation without leaving out finger types (Figure 8e), which provides an upper bound on hand CCGP. The close match between hand CCGP and decoding accuracy indicated that the laterality dimension robustly generalized across finger-types. Next, we applied cross-decoding to the finger dimension, training a classifier to discriminate between fingers of the right hand and then testing on the left hand (and vice-versa). The finger-type dimension also generalized across hands (p < 0.001), and finger-type CCGP was close to the standard decoding accuracy (Figure 8e). This result demonstrates that finger representations can be decomposed linearly into hand and finger-type building blocks.

Comparable results held for J-MC recordings, with robust decomposition of the neural code into hand and finger-type components (Supplementary Figure 9). Interestingly, J-PPC finger representations were less compositional. While above chance (p < 0.001), the finger-type CCGP (36%) was much lower than the within-hand accuracy (65%) (Supplementary Figure 10). Even when accounting for differences in neural population size, finger-type CCGP for J-PPC was lower than finger-type CCGP for N-PPC and J-MC (Supplementary Figure 11).

## Discussion

Human dexterity is characterized by our ability to quickly reach-and-grasp, as well as our ability to move individual fingers volitionally beyond basic grasp templates [7]. Such dexterity is commonly thought to be the domain of motor cortex (MC) hand knob, in part because other areas like the posterior parietal cortex (PPC) do not exhibit topographic finger activation maps like MC does [71,72]. Here, we found that neurons in two grasp-related regions of PPC are discriminative for attempted finger movements. Single-neuron tuning was robust enough for human participants to control finger BMIs using intracortical recordings. We demonstrated BMI finger control in a variety of applications, including control of a robotic hand. BMI control reached practically useful accuracies and surpassed that of a recent intracortical BMI study [27].

Our study adds to a growing number of finger BMI demonstrations. Previously, [25] demonstrated the first online neural decoding of all-five individual finger movements in human participants, using a high-density ECoG grid over the sensorimotor cortex. Similar to our study, [27] implanted intracortical arrays in a tetraplegic participant and decoded attempted finger movements using a linear classifier, achieving an offline accuracy of 67%. Recently, [24,73] achieved high-performance continuous control of flexion and extension of two finger groups. Our results contribute to prior studies by showing that simultaneous PPC+MC recordings can improve online finger decoding accuracies (Figure 6). Additionally, strong PPC-only decoding performance suggests that grasp regions of PPC (Supplementary Figure 1, [46]) may provide better dexterous finger control signals than the arm region of MC [27].

Algorithmic advances may further improve finger decoding capabilities. For example, hierarchical classifiers might be useful for classifying finger direction and finger movement [25]. Additionally, with larger data quantities or with data augmentation strategies, time-varying and nonlinear classifiers like recurrent neural networks can improve neural decoding [23,74,75]. Performance improvements may also come from decoding non-traditional variables, such as handwriting [23] or goals [10]. State-machine control (common in other assistive technologies like myoelectric prostheses [76] or Dwell) and AI-assisted hybrid control [77,78] may further improve BMI usability. In combination with somatosensory intracortical microstimulation (ICMS) to generate fingertip sensations [79,80], such methods could enable a functional hand prosthetic.

After demonstrating BMI control of the contralateral fingers, we studied representations of ipsilateral finger movements. We found that finger presses of all ten fingers were discriminable (Figure 7). At the same time, activity patterns (N-PPC and J-MC) were similar across corresponding finger movements of opposite hands (Figure 8a-b). Our results match other studies have also found shared-yet-separable hand representations in macaque anterior intraparietal area (AIP) [81] and human motor cortex [65,66]. This pattern, mixed representation of some variables over others, has previously been described as partially mixed selectivity [37], factorized representations [60], or compositional coding [58,65]. Here, the N-PPC and J-MC finger codes could be linearly decomposed into finger-type and hand building blocks (Figure 8d-e), resembling the partial compositionality described by [65] for other effectors in MC. Compositional coding has been speculated to play a number of different computational functions, from skill transfer to general cognition [37,58,59,64,65]. For neuroprosthetic applications, compositional coding reduces the amount of training data needed to calibrate decoders. Because decoders can generalize across conditions, decoders can train on only the compositional building blocks rather than every combination.

Surprisingly, J-PPC population activity did not exhibit compositionality to the same extent as N-PPC and J-MC. The difference between J-PPC and N-PPC results might stem from neuroanatomical variability [82,83] or differences in implant location. The N-PPC implant was located at the junction of the postcentral and intraparietal sulci (PC-IP), an area involved in grasping and fine finger movements [4,82,84]. PC-IP receives inputs from neighboring somatosensory cortex [83,85], suggesting that it may facilitate state estimation of the hand [70,86,87]. The J-PPC implant (Supplementary Figure 1) is in the superior parietal lobule (SPL), medial and posterior compared to the N-PPC implant. Medial and posterior areas tend to receive stronger visual inputs [83,85,88] and are more involved in reaching than grasping [89], so the recorded J-PPC population could be more involved in calculating visuomotor transforms [85,90] for visually guided reaching [83,89]. However, it is difficult to precisely compare implant regions, because the anatomical location of individual functional areas can vary widely between participants [82,83]. Future comparisons may benefit from multi-modal preoperative neuroimaging to map implant locations onto standard parcellations [91].

A follow-up question is how individual finger representations combine to construct multi-finger movements [69,92]. Previous studies of multi-effector movement have found that MC suppresses the representations of the secondary effector during dual movement of the arms or legs [65,93]. However, the representational structure of hand movements could differ fundamentally from that of other effectors [94], as human hands are uniquely dexterous and specialized [6]. fMRI studies of dual-finger movements on the same hand show that their representational structure in sensorimotor cortex follows natural hand usage patterns. These patterns could also extend to PPC recordings and to simultaneous finger movements across both hands.

## Conclusions

The posterior parietal cortex (PPC) has long been known to be involved in the reaching and grasping of objects, but less is known about its contribution to individual finger movements. Here, two tetraplegic participants controlled individual fingers through BMIs recording from the posterior parietal cortex and motor cortex (MC). Compositional coding enables decoders to both generalize across hands and discriminate between hands. Our results demonstrate that PPC and MC can provide complementary control signals for assistive neuroprosthetics.

## Data Availability

Data will be made available on the BRAIN Initiative DANDI Archive at: https://dandiarchive.org/dandiset/000252

## Acknowledgments

We thank participant N and participant J for making this research possible. We also thank Kelsie Pejsa and Viktor Scherbatyuk for administrative and technical assistance; Spencer Kellis for assistance with the robot hand.

## Supplementary Figures

**Supplementary Figure 1.**
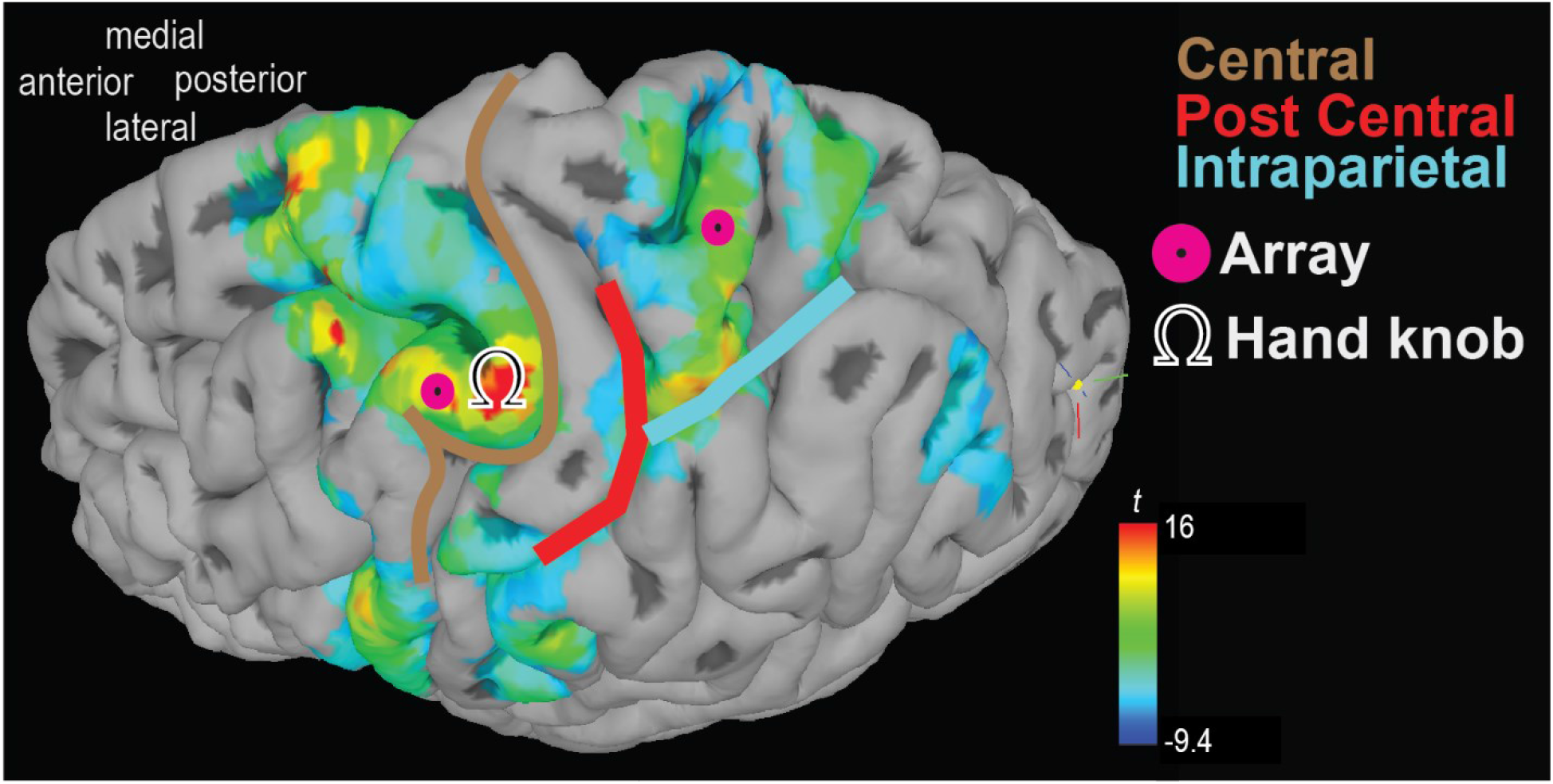
Electrode array implant locations in participant J. Microelectrode array locations overlaid on participant J’s left cerebral hemisphere. Color scale indicates fMRI activation for grasp>look/point (task described in Figure S1b of [95]). One array (denoted J-PPC) was implanted in the superior parietal lobule (SPL) of the left PPC. Another array (denoted J-MC) was implanted near the hand knob of the left motor cortex (MC).

**Supplementary Figure 2.**
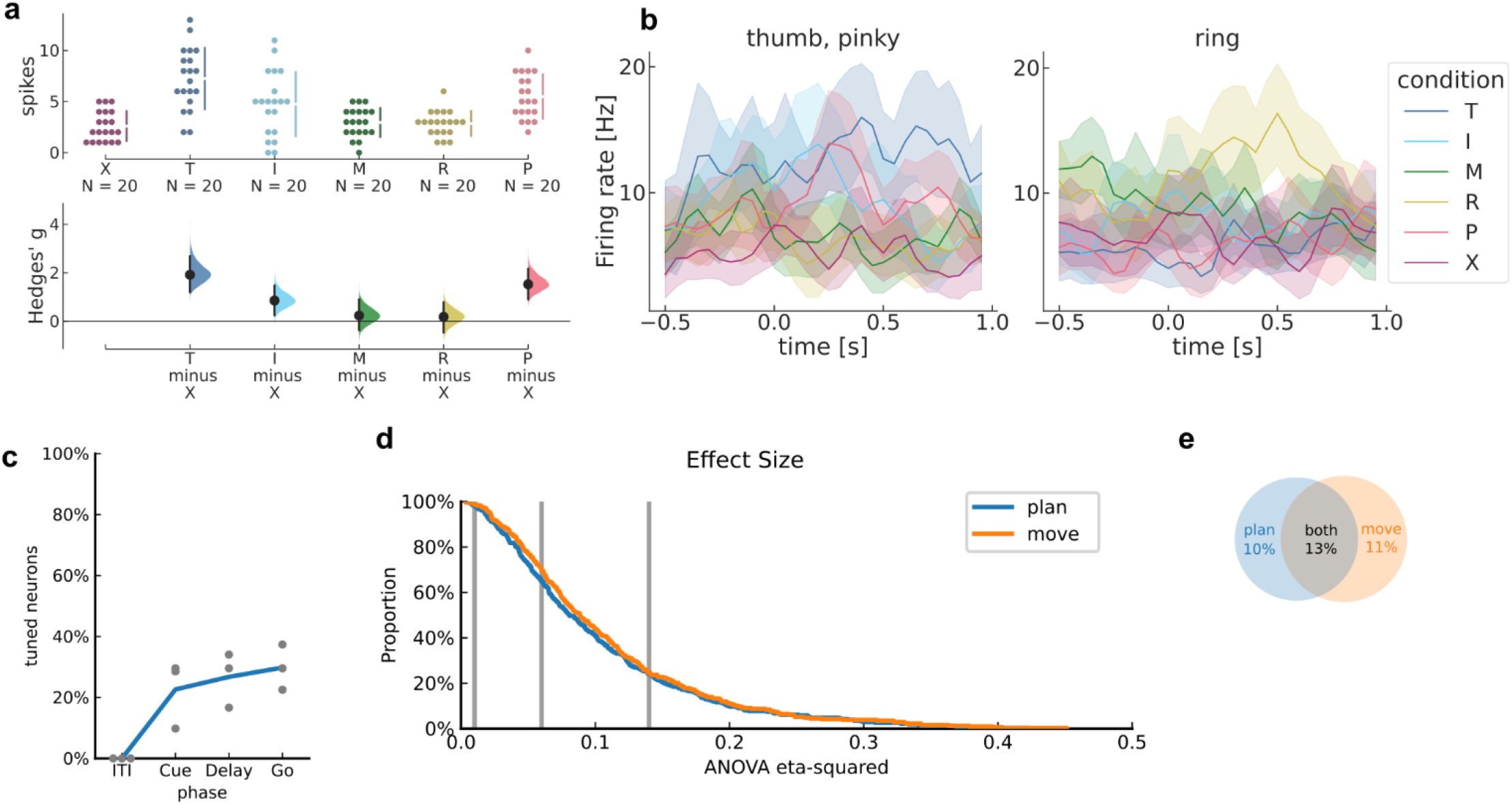
PPC single neurons discriminate between attempted finger movements (J-PPC). Similar to Figure 4 for J-PPC recordings. a) Single-trial firing rates for an example J-PPC neuron during movements of different fingers. (top) Markers correspond to the firing rate during each trial (N=120 across 6 conditions). Gapped vertical lines to the right of markers indicate +/- S.D., and each gap indicates the condition mean. (bottom) Firing rates during thumb (T), index (I), and pinky (P) presses were higher than the No-go (X) baseline. Vertical bars indicate bootstrap 95% confidence intervals (CI) of the effect size versus No-go baseline. Half-violin plots indicate bootstrap distributions. b) Mean smoothed firing rates for each finger movement for two example J-PPC neurons. Shaded areas indicate 95% CI. c) Percentage of J-PPC neurons that discriminated between finger movements in each analysis window (q < 0.05, FDR-corrected for 308 neurons). Line (blue) indicates mean across sessions. Markers (gray) indicate individual sessions. d) Complementary empirical cumulative distribution function (cECDF) visualizing the proportion of J-PPC neurons with ANOVA effect sizes (η^2^) above the corresponding x-axis value. Vertical lines (gray) indicate Cohen’s thresholds [51] for small (η^2^=0.01), medium (η^2^=0.06), and large (η^2^=0.14) effect sizes. e) Overlap of J-PPC neurons that modulated significantly (q < 0.05) with large effect sizes (η^2^ > 0.14) during movement preparation and movement execution.

**Supplementary Figure 3.**
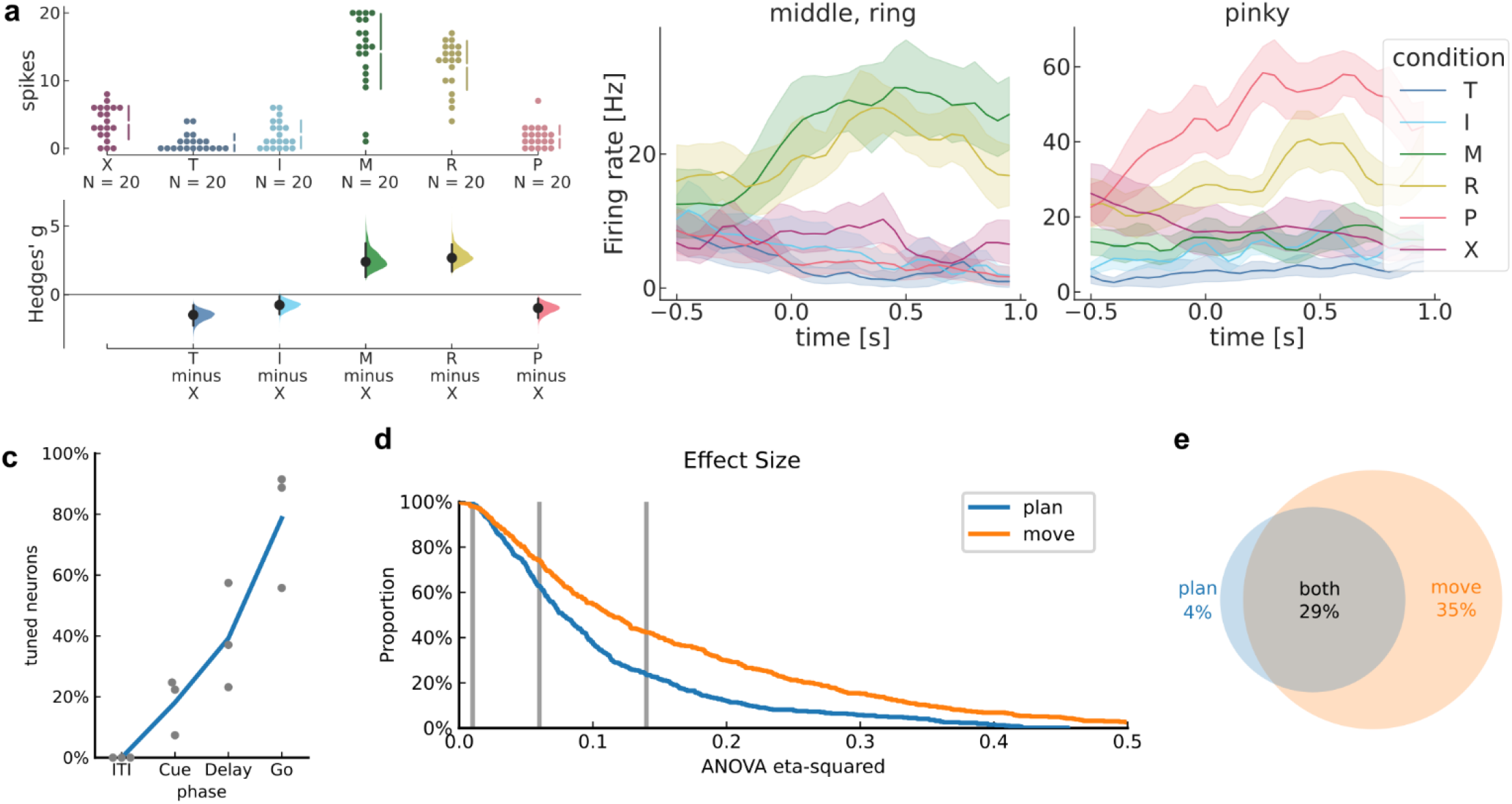
MC single neurons discriminate between attempted finger movements (J-MC). Similar to Figure 4 for J-MC recordings. a) Single-trial firing rates for an example J-MC neuron during movements of different fingers. (top) Markers correspond to the firing rate during each trial (N=120 across 6 conditions). Gapped vertical lines to the right of markers indicate +/- S.D., and each gap indicates the condition mean. (bottom) Firing rates during middle (M) and ring (R) presses were higher than the No-go (X) baseline. Vertical bars indicate bootstrap 95% confidence intervals (CI) of the effect size versus No-go baseline. Half-violin plots indicate bootstrap distributions. b) Mean smoothed firing rates for each finger movement for two example J-MC neurons. Shaded areas indicate 95% CI. c) Percentage of J-MC neurons that discriminated between finger movements in each analysis window (q < 0.05, FDR-corrected for 278 neurons). Line (blue) indicates mean across sessions. Markers (gray) indicate individual sessions. d) Complementary empirical cumulative distribution function (cECDF) visualizing the proportion of J-MC neurons with ANOVA effect sizes (η^2^) above the corresponding x-axis value. Vertical lines (gray) indicate Cohen’s thresholds [51] for small (η^2^=0.01), medium (η^2^=0.06), and large (η^2^=0.14) effect sizes. e) Overlap of J-MC neurons that modulated significantly (q < 0.05) with large effect sizes (η^2^ > 0.14) during movement preparation and movement execution.

**Supplementary Figure 4.**
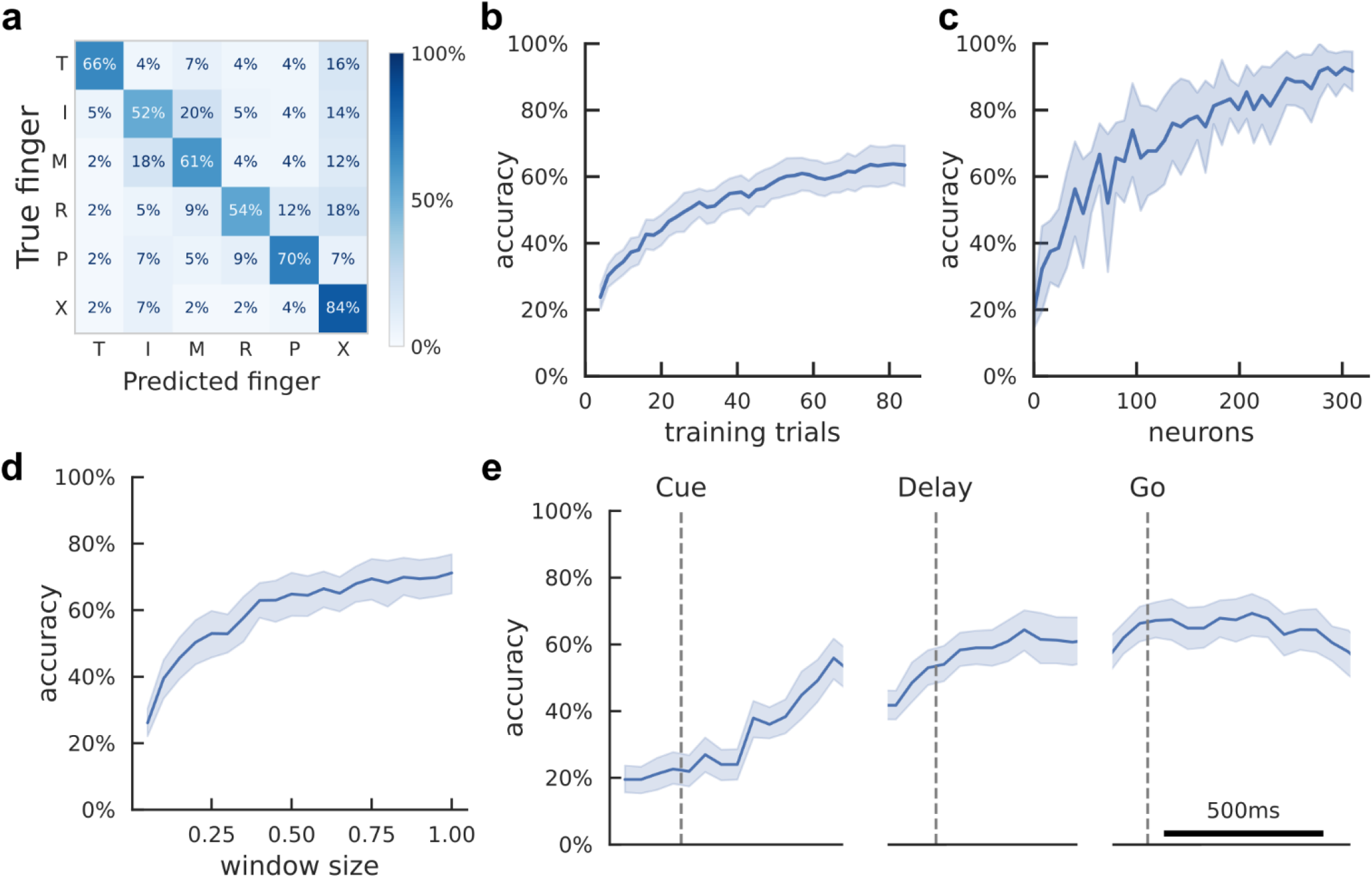
Offline classification of finger movement execution from PPC population activity (J-PPC). Similar to Figure 5 for J-PPC population recordings. a) Cross-validated confusion matrix for classifying finger movement from J-PPC neural activity. 64% accuracy, 336 trials over 3 sessions. b) Learning curve showing cross-validated accuracy as a function of the training dataset size. Shaded area indicates 95% CI over folds/sessions. c) Neuron-dropping curve (NDC) showing cross-validated accuracy as a function of recorded population size. d) Hyperparameter sweep showing cross-validated classification accuracy as a function of decode window size. Input features were the average firing rates in the window [200ms, 200ms + window size] after Go-cue. e) Cross-validated classification accuracy across the trial duration (500-ms sliding window).

**Supplementary Figure 5.**
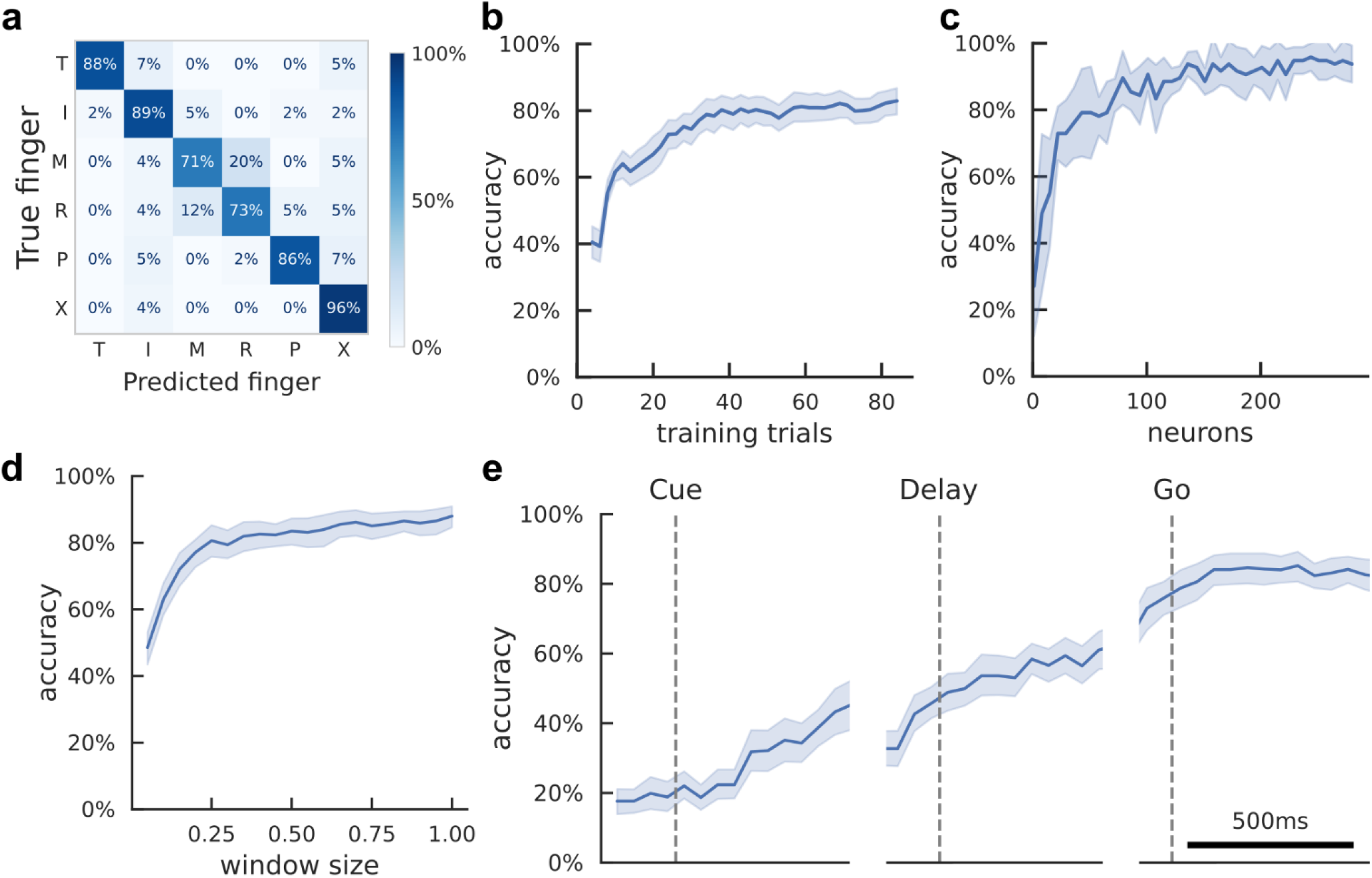
Offline classification of finger movement execution from MC population activity (J-MC). Similar to Figure 5 for J-MC population recordings. a) Cross-validated confusion matrix for classifying finger movement from J-MC neural activity. 84% accuracy, 336 trials over 3 sessions. b) Learning curve showing cross-validated accuracy as a function of the training dataset size. Shaded area indicates 95% CI over folds/sessions. c) Neuron-dropping curve (NDC) showing cross-validated accuracy as a function of recorded population size. d) Hyperparameter sweep showing cross-validated classification accuracy as a function of decode window size. Input features were the average firing rates in the window [200ms, 200ms + window size] after Go-cue. e) Cross-validated classification accuracy across the trial duration (500-ms sliding window).

**Supplementary Figure 6.**
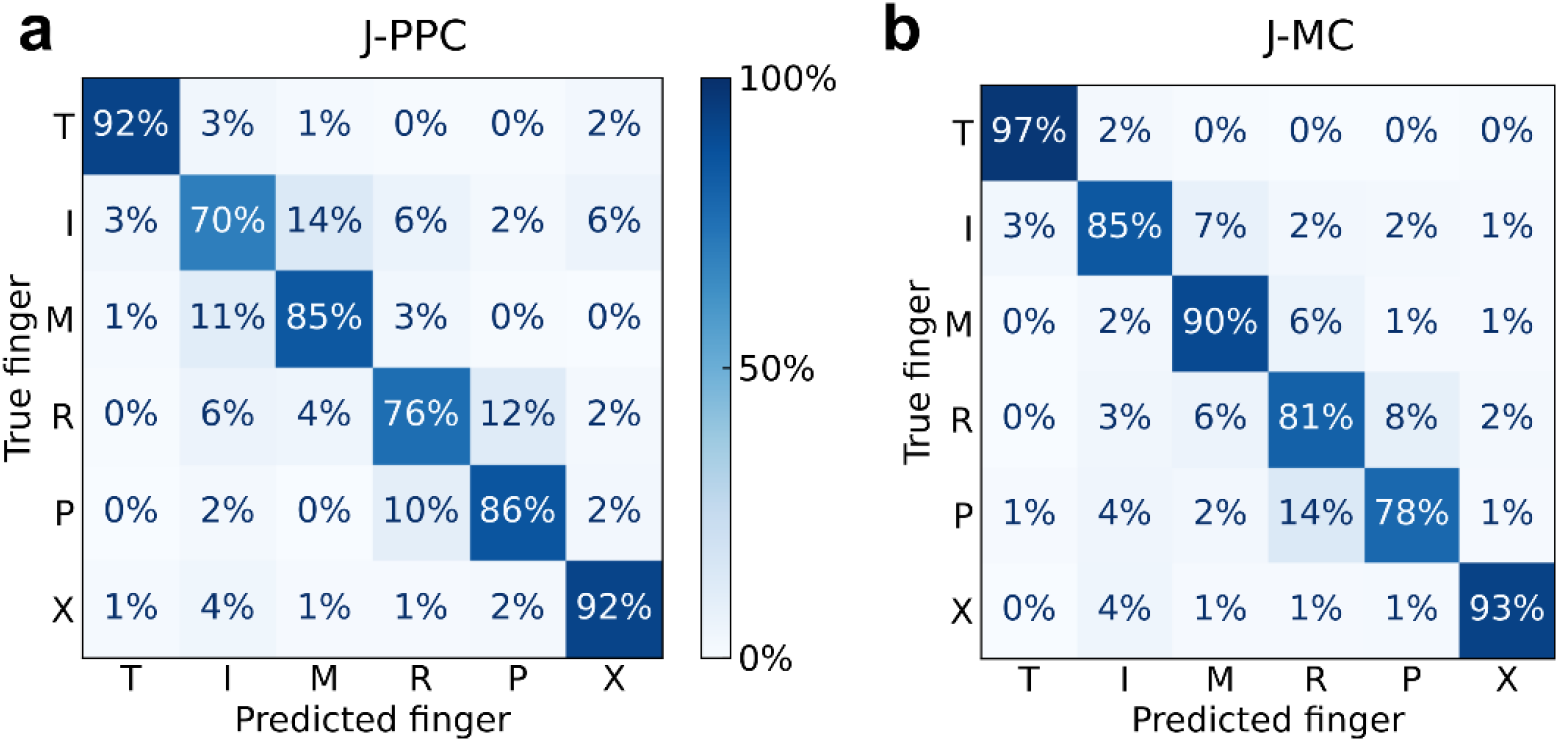
Retrospective BMI accuracy when decoding from a single electrode array. a) Offline approximation of single-array BMI accuracy. We trained the linear classifier on earlier run-blocks [0, 1, …, M-1] and evaluated on run-block M, repeating for all run-blocks in each session. 83% +/- 7% accuracy (chance 17%), 1488 trials over 8 sessions. b) Same as (a) using J-MC population activity. 87% +/- 8% accuracy (chance 17%), 1488 trials over 8 sessions.

**Supplementary Figure 7.**
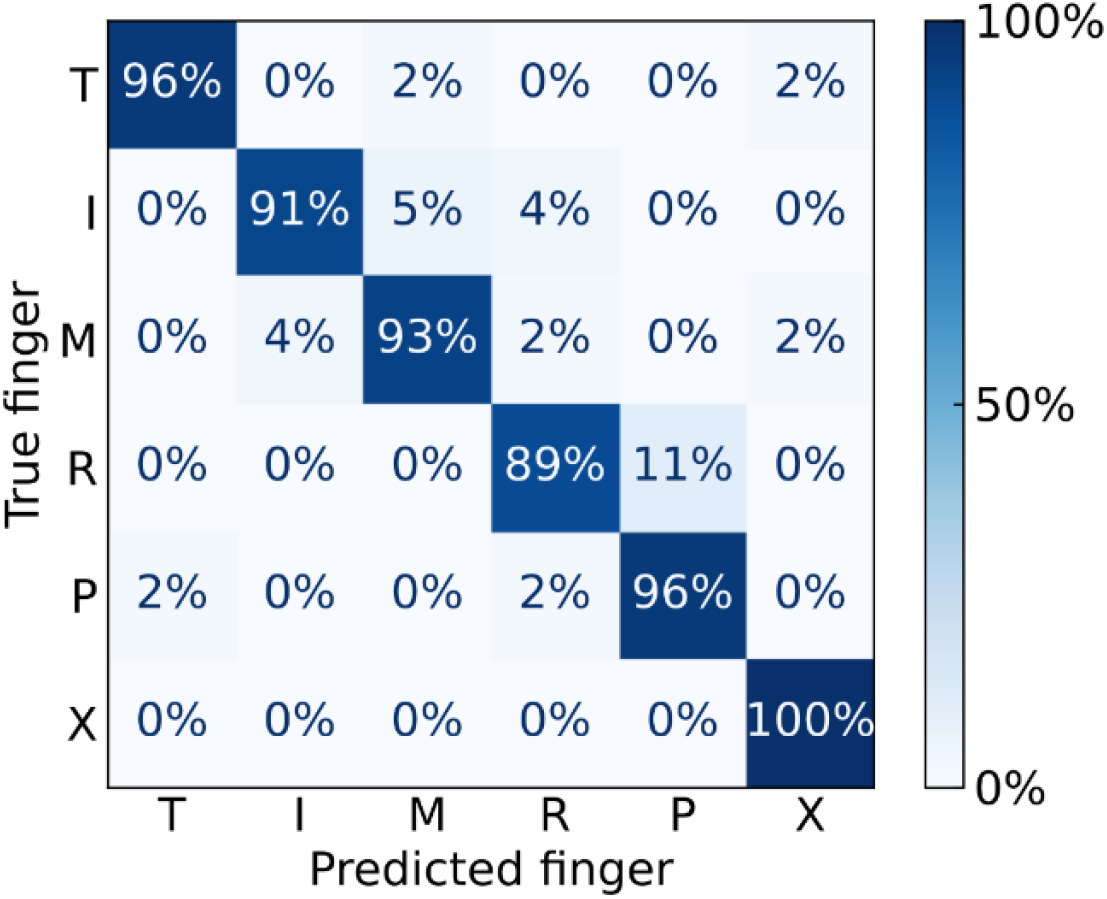
Left-hand BMI classification of individual finger movements. Confusion matrix for participant J (PPC+MC), left-hand finger presses. 94% accuracy, 336 trials over 3 sessions.

**Supplementary Figure 8.**
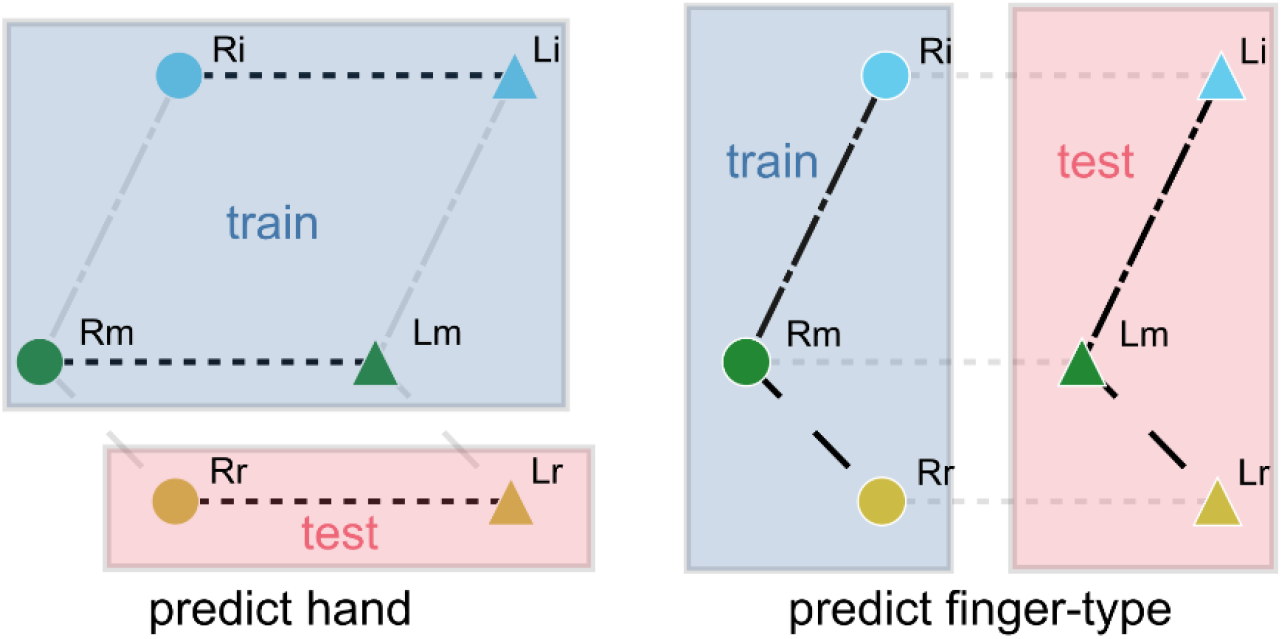
Cross-condition generalization paradigm to assess compositional coding hypothesis. Compositional coding should enable a linear decoder to generalize to unseen test conditions. (left) A left-vs-right-hand decoder was trained on 4 pairs of fingers, then evaluated on the held-out finger-type. Cross-condition generalization accuracies (CCGP) were averaged over all folds of hold-one-finger-type-out. (right) A 5-class finger-type decoder was trained on the right hand and evaluated on the left hand (and vice versa). (both) Plot only shows markers for the right and left index, middle, and ring fingers, but the actual procedure included all ten fingers.

**Supplementary Figure 9.**
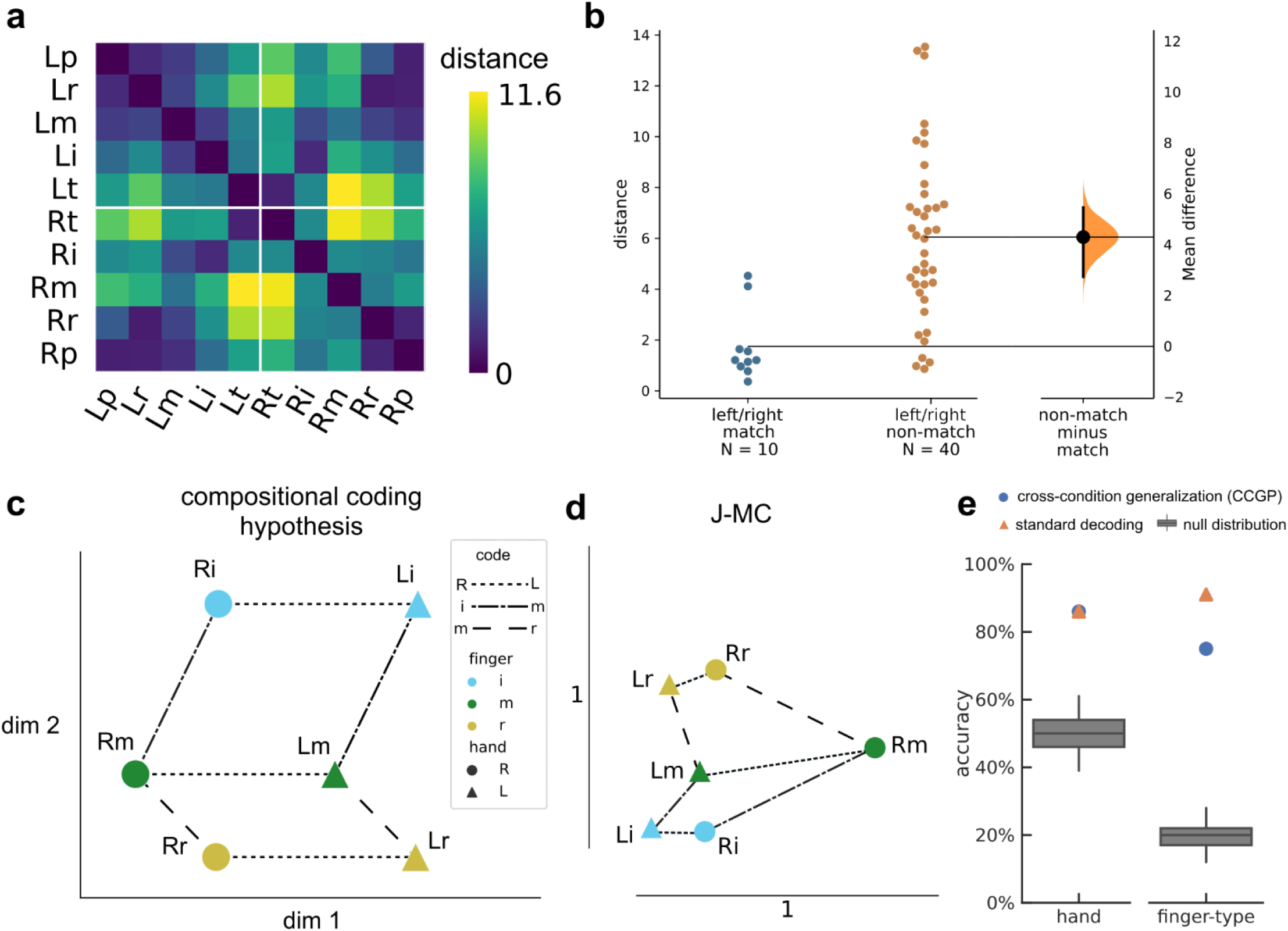
Representational geometry of contralateral and ipsilateral finger movements (J-MC) Similar to Figure 8 for J-MC population recordings. a) Cross-validated squared Mahalanobis distances between J-MC activity patterns during the contralateral/ipsilateral finger press task. Distances were averaged over the 2 sessions. b) Non-matching finger pairs have larger distances than matching finger pairs. Each circle is one pairwise distance, aggregated across 2 sessions. c) Example schematic of perfect compositionality along hand and finger-type components. Line styles indicate groups of parallel, identical vectors. A compositional code can be represented as the linear summation of simple building blocks. For example, the Rm population activity can be constructed from the summation: Li + left->right + index->middle. For visual clarity, only three finger-types (index, middle, ring) are shown. d) Representational geometry of finger movements corresponding to J-MC distances (a), visualized in 2-D using multidimensional scaling (MDS). We used Generalized Procrustes analysis (with scaling) to align across 2 sessions. Markers indicate mean across sessions. Scale bars shown. e) Linear decoders generalized (Supplementary Figure 8) across finger-type to classify hand (left) and across hand to classify finger-type (right) (p < 0.001, permutation test), indicating that movement representations were compositional across finger-type and hand dimensions.

**Supplementary Figure 10.**
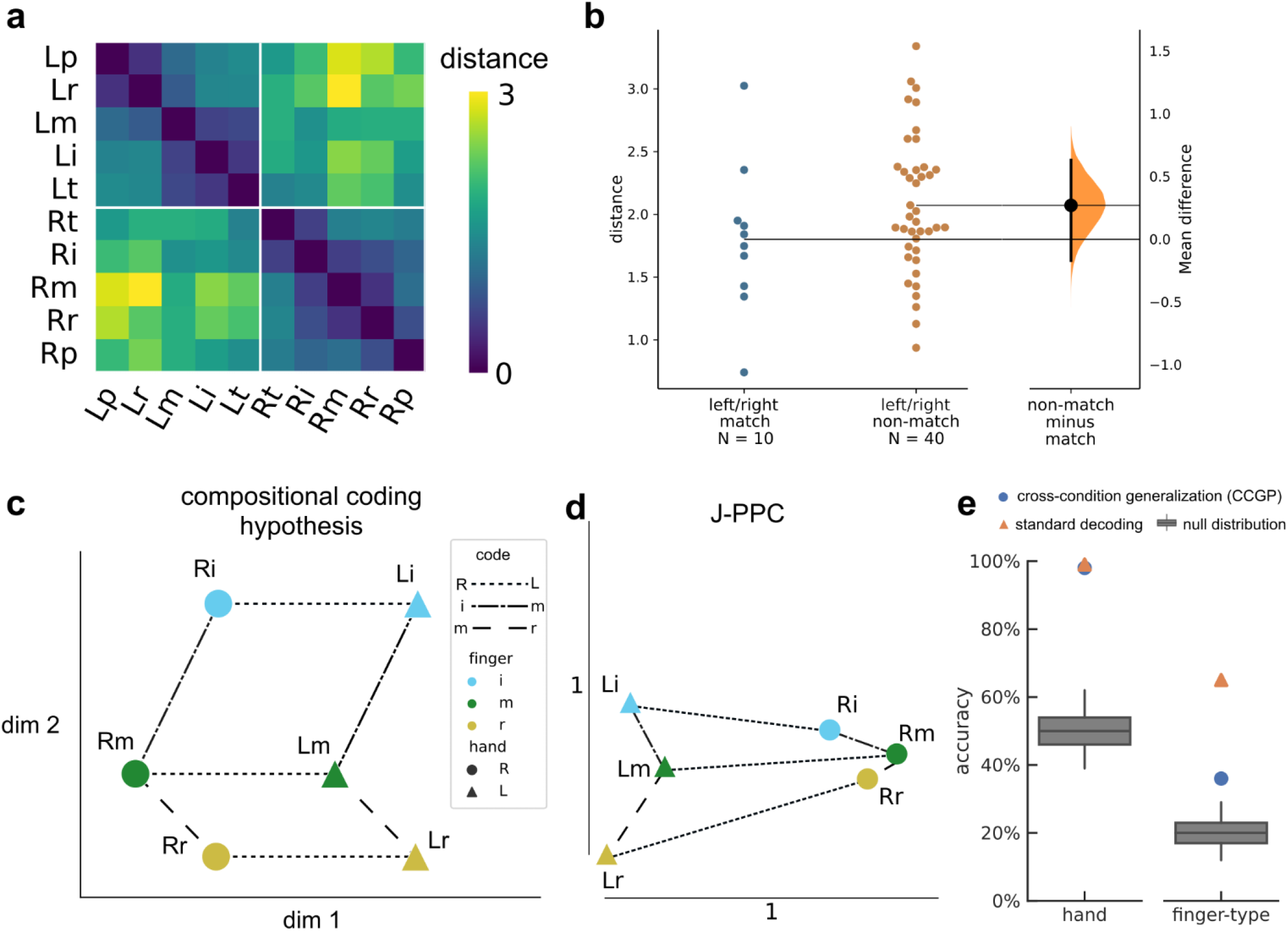
Representational geometry of contralateral and ipsilateral finger movements (J-PPC) Similar to Figure 8 for J-PPC population recordings. a) Cross-validated squared Mahalanobis distances between J-PPC activity patterns during the contralateral/ipsilateral finger press task. Distances were averaged over the 2 sessions. b) Comparison of distances between matching finger pairs and non-matching finger pairs. Each circle is one pairwise distance, aggregated across 2 sessions. c) Example schematic of perfect compositionality along hand and finger-type components. Line styles indicate groups of parallel, identical vectors. A compositional code can be represented as the linear summation of simple building blocks. For example, the Rm population activity can be constructed from the summation: Li + left->right + index->middle. For visual clarity, only three finger-types (index, middle, ring) are shown. d) Representational geometry of finger movements corresponding to J-PPC distances (a), visualized in 2-D using multidimensional scaling (MDS). We used Generalized Procrustes analysis (with scaling) to align across 2 sessions. Markers indicate mean across sessions. Scale bars shown. e) (left) Linear decoders generalized (Supplementary Figure 8) across finger-type to classify hand (left; p < 0.001, permutation test). (right) Finger-type cross-condition generalization performance (CCGP; blue circle) was lower than standard decoding accuracy (orange triangle).

**Supplementary Figure 11.**
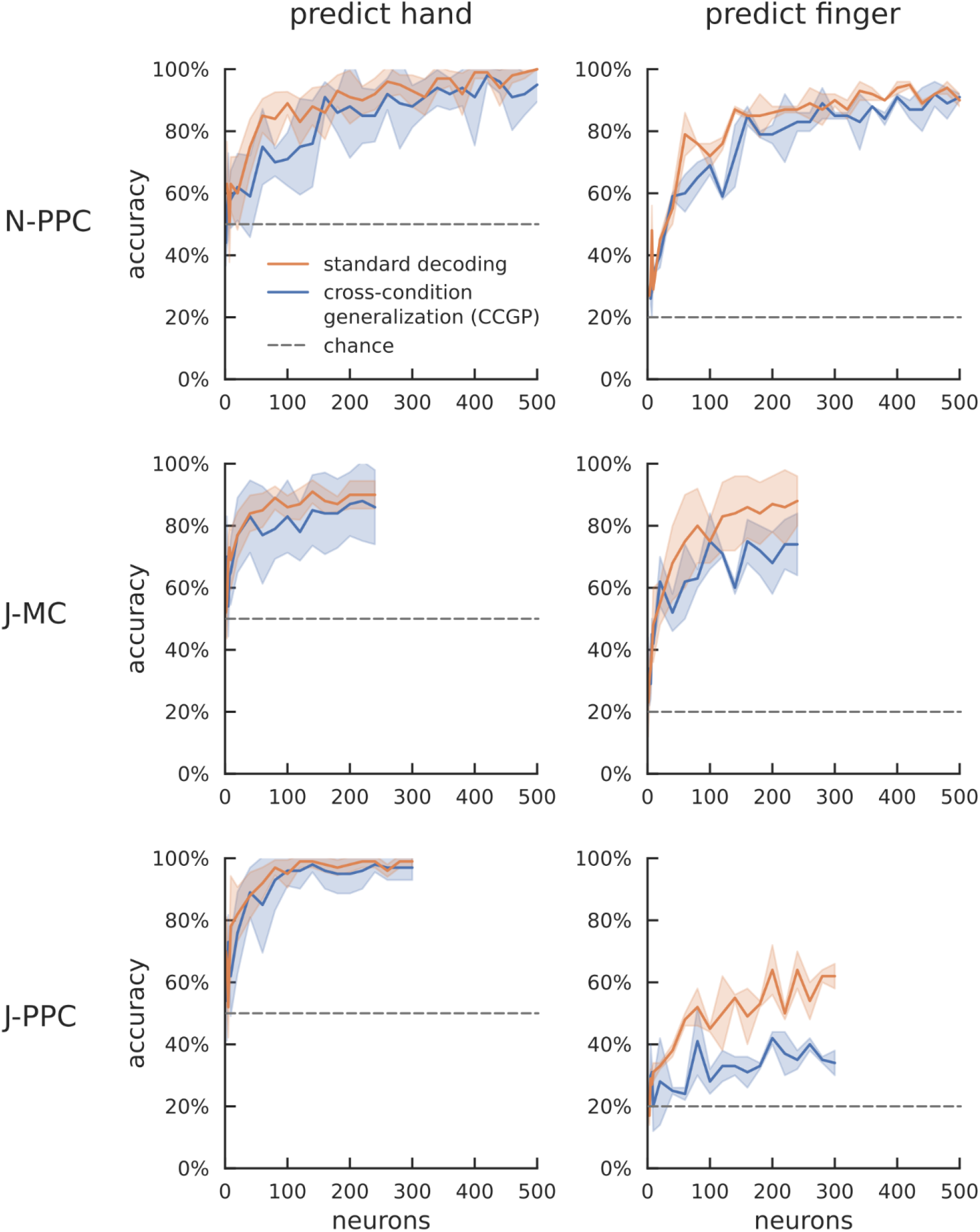
Neuron-dropping curve for cross-condition generalization performance (CCGP). Neuron dropping curves show generalization accuracy as a function of neural population size. Panel columns correspond to the variable predicted (left: hand; right: finger-type). Panel rows correspond to the recorded neural population. Hand CCGP matched standard decoding accuracy in all recorded neural populations, but finger-type decoders trained on J-PPC activity did not generalize well across hands (bottom right panel). 1111 N-PPC neurons were recorded across 10 sessions, but we truncated the dropping curve at 500 neurons to match scales across participants.

**Supplementary Table 1.** Phase durations for each task. Table of phase durations for the different tasks and participants. Blank cells indicate that the task did not include that phase. Ranges indicate a uniform-random distribution per trial.

| Task | Participant | Phase |  |  |  |  |  |
| --- | --- | --- | --- | --- | --- | --- | --- |
|  |  | Pre-Cue | Cue | Delay | Go | ITI | Feedback |
| Alternating Cues | N | 1 sec | 0.75 sec | 1.25 - 2 sec | 1 sec | 2 sec |  |
| Alternating Cues | J | 0.5 sec | 1.5 sec | 0.5 - 1 sec | 1 sec | 0.5 - 1 sec |  |
| Reaction time,<br>randomized<br>location | N and J |  |  |  | 1.5 sec |  | 1.5 sec |
| Ten-finger | N |  | 1.5 or 1 sec | 1 or 1-1.5<br>sec | 1 sec | 2 or 1.5 sec |  |
| Ten-finger | J |  | 1 or 1.5 sec | 1-1.5 or<br>0.75-1 sec | 1 or 2 sec | 1.5 or 2 sec |  |

**Supplementary Table 2.**
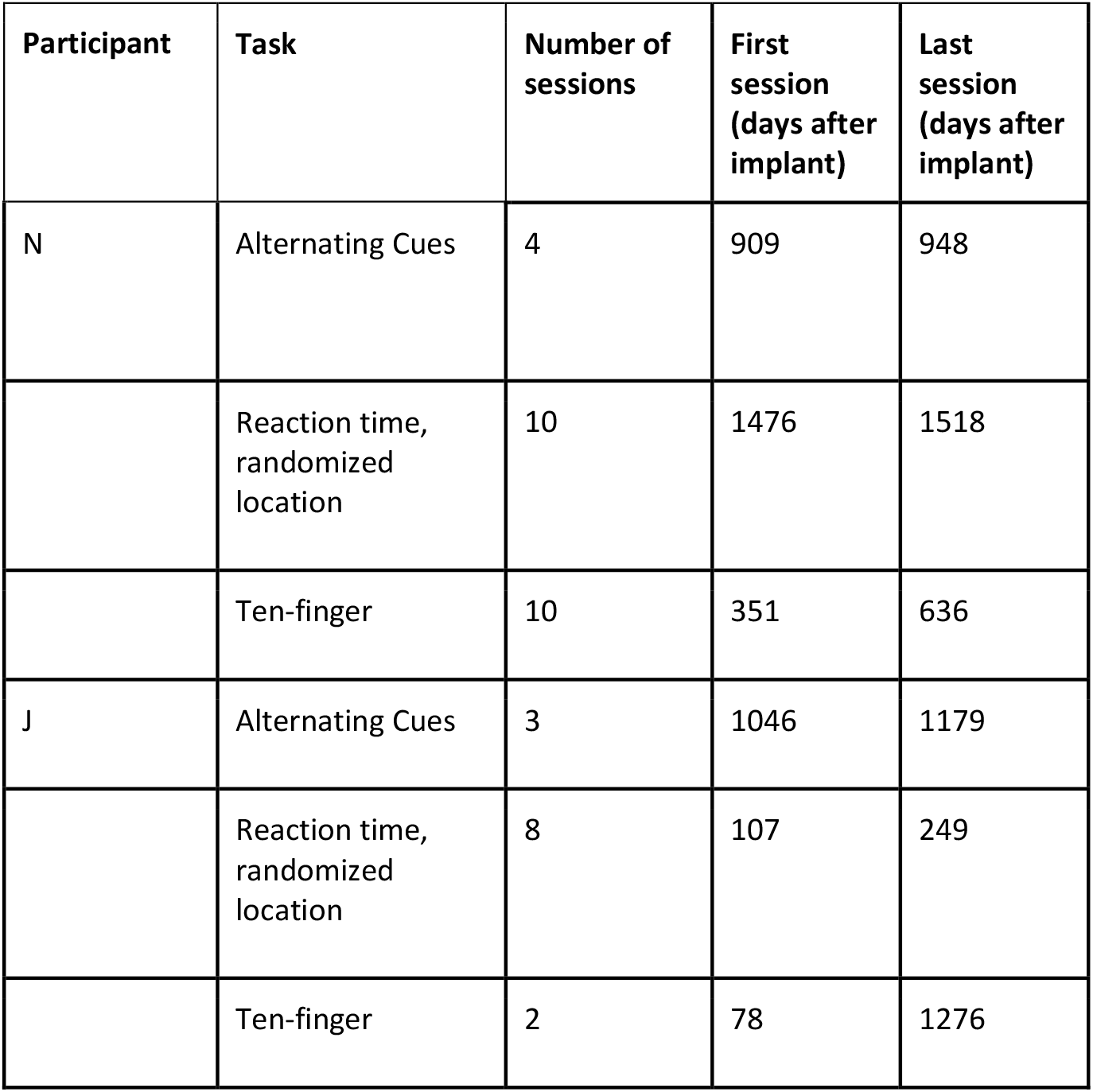
Date ranges for each task. Dates of first and last session for each task. Although the Alternating Cues data was presented first in the Results section, this data was collected from participant J later than the reaction-time data. The drop in neural yield between these dates affected classification performance.

| Participant | Task | Number of sessions | First session (days after implant) | Last session (days after implant) |
| --- | --- | --- | --- | --- |
| N | Alternating Cues | 4 | 909 | 948 |
|  | Reaction time, randomized location | 10 | 1476 | 1518 |
|  | Ten-finger | 10 | 351 | 636 |
| J | Alternating Cues | 3 | 1046 | 1179 |
|  | Reaction time, randomized location | 8 | 107 | 249 |
|  | Ten-finger | 2 | 78 | 1276 |

## Notes

### Competing Interest Statement

The authors have declared no competing interest.

### Clinical Trial

NCT01958086

### Funding Statement

This study was funded by NIH National Eye Institute grants UG1EY032039 and R01EY015545, the T&C Chen Brain-machine Interface Center (C.G., T.A., K.K., J.G., R.A.A.), Amazon AI4Science Fellowship (C.G.), and the Boswell Foundation (R.A.A).

### Author Declarations

IRB of California Institute of Technology gave ethical approval for this work. IRB of Casa Colina Hospital and Centers for Healthcare gave ethical approval for this work. IRB of University of California Los Angeles gave ethical approval for this work.

